# A NOVEL METHOD FOR HANDLING PRE-EXISTING CONDITIONS IN PREDICTION MODELS FOR COVID-19 DEATH

**DOI:** 10.1101/2022.01.22.22269694

**Authors:** Glen H. Murata, Allison E. Murata, Heather M. Campbell, Benjamin H. Mcmahon, Jenny T. Mao

## Abstract

**Objective:** To derive a predicted probability of death (PDeathDx) based upon complete sets of ICD-10 codes assigned to patients prior to their diagnosis of COVID-19. PDeathDx is intended for use as a summary metric for pre-existing conditions in multivariate models for COVID-19 death.

**Methods:** Cases were identified through the COVID-19 Shared Data Resource (CSDR) of the Department of Veterans Affairs. The diagnosis required at least one positive nucleic acid amplification test (NAAT). The primary outcome was death within 60 days of the first positive test. We retrieved all diagnoses entered into the electronic medical record for visits, on problem lists, and at the time of hospital discharge if they were at least 14 days prior to the NAAT. ICD-9 codes were converted to ICD-10 equivalents using a crosswalk provided by the Centers for Medicare/Medicaid Services. ICD-10 codes were converted to their category diagnoses defined as all columns to the left of the decimal point. Each patient was considered to have or not have each category diagnosis prior to the NAAT. A computer program calculated the number of cases for each category diagnosis, the relative risk (RR) of death, and its confidence interval (CI) using a Bonferroni adjustment for multiple comparisons. RRs were re-centered by subtracting 1 so that high-risk conditions had a positive value while protective conditions had a negative one. Diagnoses found to be significant were entered into a logistic model for death in a stepwise fashion. Each patient was assigned (RR-1) to each category diagnosis if they had the condition or 0 otherwise. The resulting model was used to derive PDeathDx for each patient and the area under its receiver operating characteristic (ROC) curve calculated. Single variable logistic models were also derived for age at diagnosis, the Charlson 2-year (Charl2Yr) and lifetime (CharlEver) scores, and the Elixhauser 2-year (Elix2Yrs) and lifetime (ElixEver) scores. Stata was used to compare the ROCs for PDeathDx and each of the other metrics.

**Results:** On September 30, 2021 there were 347,220 COVID-19 patients in the CSDR. 18,120 patients (5.33%) died within 60 days of their diagnosis. After consolidating ICD-9 and ICD-10 codes, 29,162,710 separate diagnoses were given to the subjects representing 41,341 ICD-10 codes. This set was reduced to 1,890 category diagnoses assigned to the group for the first time on 19,184,437 occasions. Of the 1,890 category diagnoses, 425 involved >= 100 subjects and had a lower boundary for the CI >= 1.50 (a high-risk condition) or upper boundary <= 0.80 (a protective condition). Stepwise logistic regression showed that 153 were statistically significant, independent predictors of death. PDeathDx was slightly less powerful than age as a discriminator (ROC = 0.811 +/- 0.002 vs 0.812 +/- 0.001, respectively; P < 0.001) but was superior to the Charl2Yr (ROC = 0.727 +/- 0.002; P < 0.001), CharlEver (ROC = 0.753 +/- 0.002; P <= 0.001), Elix2Yr (ROC = 0.694 +/- 0.002; P < 0.001); and ElixEver (ROC = 0.731 +/- 0.002; P < 0.001). Univariate analysis and multivariate modeling showed that many of the most high-risk conditions are under-represented or not included in the Charlson Index. These include hypertension, dementia, degenerative neurologic disease, or diagnoses associated with severe physical disability.

**Conclusions:** Our method for handling pre-existing conditions in multivariate analysis has many advantages over conventional comorbidity indices. The approach can be applied to any condition or outcome, can use any categorical predictors including medications, creates its own condition weights, handles rare as well as protective conditions, and returns actionable information to providers. The latter include the specific ICD-10 groups, their contribution to the risk, and their rank order of importance. Finally, PDeathDx is equivalent to age as a discriminator of outcomes and outperforms 4 other comorbidity scores. If validated by others, this approach provides an alternative and more robust approach to handling comorbidities in multivariate models.

## INTRODUCTION

Many mathematical models have been proposed for predicting death from COVID-19 infection (1-5). They have gained popularity because they are very useful for managing patients and allocating scarce resources. One of the most important components of these models is the set of pre-existing conditions. Many diseases increase the mortality rate because they diminish the host response to infection, cause end-organ dysfunction that is further compromised by COVID-19, or severely limit life expectancy and functional status. One method for handling these conditions is to gather them under broad categories (“malignancy”) and derive a regression coefficient for each grouping (3, 4). Another approach is to use a comorbidity scoring system such as the Charlson Comorbidity Index (CCI) or Elixhauser (ELIX) score (1, 2, 5). With this approach, the groupings are assigned weights, and the weights are added to generate a summary score. The summary score is then used as a covariate in the model. CCI and ELIX have been extensively validated and applied to many conditions (6). Moreover, mathematical modeling has shown that they provide all that is necessary to handle confounding from the underlying diagnoses (7). However, there are still problems with either approach:

1. CCI was developed to predict mortality one year after hospitalization. It is less clear that the same conditions are the most influential for outcomes mediated by an immune response such as breakthrough infections.
2. Rare diseases are not well represented. As a result, a person with a common disease of limited impact could be given a poorer prognosis than another with a rare but lethal condition.
3. There is often little evidence that members of each group have the same prognostic significance.
4. Neither method handles conditions that are protective.
5. These approaches do not return actionable information to clinicians. While they may become aware of their patients’ risk, providers are not informed of the sources of such risk. As a result, they cannot take steps to mitigate that risk through further studies or treatments.

A more sophisticated approach is to do a systematic survey of all pre-existing conditions, determine which have an impact on outcomes, and generate a predicted probability of death that represents the aggregate risk posed by those that are statistically significant. Like a propensity score, this predicted probability of death (PDeathDx) can be used as a variable in the model. Fortunately, advanced computer methods have made it possible to generate these estimates. The purpose of this study is to describe a novel approach to handling comorbidities in COVID-19 prediction models and compare the performance of PDeathDx with conventional comorbidity measures in distinguishing fatal from non-fatal cases.

## METHODS

Cases were identified through VA’s COVID-19 Shared Data Resource (CSDR). Membership in this registry requires at least one positive nucleic acid amplification test (NAAT). The primary outcome was death within 60 days of the diagnosis. The outcome was retrieved from the post-index conditions file of the CSDR, which assigns a 1 to those who died and 0 otherwise. Likewise, the 2-year CCI score (Charl2Yrs), lifetime CCI score (CharlEver), 2-year ELIX score (Elix2Yrs), and lifetime ELIX score (ElixEver) were retrieved from CSDR for each patient. Pre-existing conditions were identified by reviewing all diagnoses entered into the electronic medical record for outpatient visits, on patient problem lists, or at the time of hospital discharge. “Pre-existing” refers to entries made at least 14 days prior to the diagnosis of COVID-19. This precaution excludes any entries that may have been made during the pre-symptomatic phases of COVID-19. ICD-9 codes were converted to ICD-10 using a crosswalk provided by the Centers for Medicare/Medicaid Services. A “category diagnosis” was defined as all characters preceding the decimal point for ICD-10 codes or the ICD-9 equivalent. A patient was considered to have or not have each category condition prior to the COVID diagnosis. A computer program was used to identify all patients with a given condition who died or survived as well as all patients without the condition who died or survived. The software used these cell frequencies to derive the relative risk of death associated with the condition (RR) and its confidence interval (CI). CI were adjusted for multiple comparisons by the Bonferroni method. A category diagnosis was considered to have a significant effect on the outcome if there were at least 100 cases and if the lower limit for the CI was >= 1.5 or the upper limit was <= 0.80. Another software program was used to create a diagnostic grid in which each patient was assigned a value for all significant category diagnoses. Recall that a condition with a relative risk of 1 has no effect on the outcome. The scale for relative risk was therefore centered on 1 by subtracting 1 from the relative risk and entering that value for the corresponding category condition if present. Those without the condition were treated as if they had a condition with no prognostic significance and assigned a value of 0. The diagnostic grid was exported to a statistical program (Stata). Stepwise logistic regression was used to identify those that were independently predictive of death. This procedure adjusted the contribution of each condition for the presence of the others, assigned a predicted probability of death to each patient (PDeathDx), and used PDeathDx to generate an area under a Receiver Operating Characteristic curve (ROC area). The product of coefficient * (RR – 1) is the contribution of each condition to the logit function if present. It was therefore used to rank order the importance of the category diagnoses. Differences in categorical variables were tested by chi-square analysis; differences in continuous variables were analyzed by the unpaired student’s t-test or rank sum test. Separate logistic models were used to examine the effect of age, Charl2Yrs, CharlEver, Elix2Yrs, and ElixEver as single predictors of death. Stata provides a function that compares the ROC areas for pairs of these variables.

## RESULTS

On September 30, 2021, there were 347,220 COVID-19 patients in VA’s COVID-19 Shared Data Resource. 339,772 (or 97.9%) had at least one pre-existing condition and form the basis for this report. The mean age at the time of diagnosis was 58.6 ± 16.7 years; 84.1% were male; 22.9% were members of a racial minority; 9.0% were Hispanic; 95.8% were veterans; 0.7% were on supplemental oxygen; and 11.8% were current smokers. 9.1% had been fully vaccinated at least 14 days prior to the COVID-19 diagnosis. 21.5% acquired their infections after July 1, 2021 and were presumed to have the delta variant. Overall, 18,120 patients (5.33%) died within 60 days of their diagnosis.

For the study cohort, 82,578,233 ICD-9 codes had been entered into the electronic record before the VA converted to ICD-10 codes in 2015. Of these, 81,671,483 (or 98.9%) were successfully converted to an ICD-10 equivalent using the CMS crosswalk. In addition, 78,976,269 ICD-10 codes were entered at least 14 days prior to the COVID-19 diagnosis. After consolidation, 29,162,710 separate diagnoses were given to the subjects representing 41,341 ICD-10 codes. The sample size was insufficient to test each ICD-10 code for its prognostic significance. The individual codes were therefore reduced to 1,890 category diagnoses which were assigned to the group for the first time on 19,184,437 occasions.

Of the 1,890 category diagnoses, 425 involved ≥ 100 subjects and had a lower boundary for the CI >= 1.50 or upper boundary <= 0.80 (Table 1). One diagnosis (Z11: screening for other viral diseases) was given to 120,308 subjects and had a high RR for death; it was removed because it may have been used for COVID-19 testing before there was a suitable ICD-10 code. Preliminary regressions indicated that another 26 provided a perfect prediction of the outcome when present, while 20 were affected by collinearity. A diagnostic grid was therefore assembled where each patient was assigned a value for each of the remaining 378 category diagnoses. If the diagnosis was present, (RR – 1) was assigned as its value and 0 otherwise. Stepwise logistic regression showed that 153 were statistically significant, independent predictors of death (Table 2). PDeathDx was derived for each patient and its ROC area determined. This analysis showed that PDeathDx provided excellent discrimination between those who died and those who survived (ROC area = 0.811 ± 0.002). Single variable logistic models were also constructed for age at diagnosis, Charl2Yrs, CharlEver, Elix2Yrs, and ElixEver, and ROC areas determined for their predicted probabilities. PDeathDx was less powerful than age as a discriminator (ROC = 0.812 ± 0.001; P < 0.001) but was superior to the Charl2Yr (ROC = 0.727 ± 0.002; P < 0.001), CharlEver (ROC = 0.753 ± 0.002; P <= 0.001), Elix2Yr (ROC = 0.694 ± 0.002; P < 0.001); and ElixEver (ROC = 0.731 ± 0.002; P < 0.001).

**TABLE 1:** ICD-10 CATEGORY DIAGNOSES PREDICTIVE OF DEATH ON UNIVARIATE ANALYSIS. * Most Common ICD-10 Code and Description for Each Category Diagnosis

| Root ICD-10 | ICD-10 Code* | ICD-10 Description* | No. of Cases | Case Rate | Control Rate | Relative Risk | Lower CI | Higher CI |
| --- | --- | --- | --- | --- | --- | --- | --- | --- |
| G30 | G30.9 | Alzheimer's disease, unspecified | 4348 | 0.2672 | 0.0506 | **5.2861** | 4.7347 | 5.9017 |
| F03 | F03.90 | Unspecified dementia without behavioral disturbance | 13895 | 0.2340 | 0.0456 | **5.1278** | 4.7678 | 5.5150 |
| F02 | F02.80 | Dementia in oth diseases classd elswhr w/o behavrl disturb | 8684 | 0.2379 | 0.0485 | **4.9065** | 4.4977 | 5.3524 |
| I10 | I10. | Essential (primary) hypertension | 208817 | 0.0768 | 0.0159 | **4.8387** | 4.3920 | 5.3309 |
| F01 | F01.50 | Vascular dementia without behavioral disturbance | 6033 | 0.2342 | 0.0501 | **4.6786** | 4.2212 | 5.1856 |
| Z66 | Z66. | Do not resuscitate | 6448 | 0.2244 | 0.0500 | **4.4864** | 4.0498 | 4.9700 |
| R62 | R62.7 | Adult failure to thrive | 3414 | 0.2209 | 0.0516 | **4.2777** | 3.7238 | 4.9140 |
| R54 | R54. | Age-related physical debility | 11357 | 0.1983 | 0.0483 | **4.1040** | 3.7668 | 4.4714 |
| C93 | C93.10 | Chronic myelomonocytic leukemia not achieve remission | 122 | 0.2131 | 0.0533 | **4.0005** | 1.9240 | 8.3180 |
| D46 | D46.9 | Myelodysplastic syndrome, unspecified | 755 | 0.2106 | 0.0530 | **3.9750** | 2.9513 | 5.3539 |
| L89 | L89.90 | Pressure ulcer of unspecified site, unspecified stage | 8092 | 0.1908 | 0.0500 | **3.8180** | 3.4499 | 4.2253 |
| I13 | I13.0 | Hyp hrt & chr kdny dis w hrt fail and stg 1-4/unsp chr kdny | 9823 | 0.1844 | 0.0494 | **3.7299** | 3.3925 | 4.1008 |
| J64 | J64. | Unspecified pneumoconiosis | 107 | 0.1963 | 0.0533 | **3.6833** | 1.6173 | 8.3882 |
| C95 | C95.90 | Leukemia, unspecified not having achieved remission | 609 | 0.1938 | 0.0531 | **3.6505** | 2.5754 | 5.1743 |
| R64 | R64. | Cachexia | 797 | 0.1932 | 0.0530 | **3.6457** | 2.6851 | 4.9499 |
| E43 | E43. | Unspecified severe protein-calorie malnutrition | 1597 | 0.1904 | 0.0527 | **3.6133** | 2.9023 | 4.4984 |
| Z74 | Z74.09 | Other reduced mobility | 17940 | 0.1681 | 0.0469 | **3.5822** | 3.3154 | 3.8704 |
| D63 | D63.1 | Anemia in chronic kidney disease | 14823 | 0.1703 | 0.0480 | **3.5494** | 3.2667 | 3.8565 |
| C24 | C24.1 | Malignant neoplasm of ampulla of Vater | 122 | 0.1885 | 0.0533 | **3.5383** | 1.6054 | 7.7983 |
| F05 | F05. | Delirium due to known physiological condition | 3612 | 0.1822 | 0.0519 | **3.5070** | 3.0142 | 4.0803 |
| Y95 | Y95. | Nosocomial condition | 355 | 0.1859 | 0.0532 | **3.4952** | 2.1891 | 5.5806 |
| W06 | W06.XX<br>XA | Fall from bed, initial encounter | 2167 | 0.1832 | 0.0525 | **3.4898** | 2.8768 | 4.2334 |
| C78 | C78.7 | Secondary malig neoplasm of liver and intrahepatic bile duct | 1305 | 0.1839 | 0.0528 | **3.4814** | 2.7193 | 4.4570 |
| J90 | J90. | Pleural effusion, not elsewhere classified | 5673 | 0.1754 | 0.0513 | **3.4218** | 3.0197 | 3.8774 |
| J69 | J69.0 | Pneumonitis due to inhalation of food and vomit | 2896 | 0.1785 | 0.0523 | **3.4165** | 2.8812 | 4.0511 |
| I50 | I50.9 | Heart failure, unspecified | 36427 | 0.1447 | 0.0424 | **3.4162** | 3.2021 | 3.6445 |
| N18 | N18.9 | Chronic kidney disease, unspecified | 48428 | 0.1352 | 0.0397 | **3.4042** | 3.2006 | 3.6206 |
| I75 | I75.029 | Atheroembolism of unspecified lower extremity | 138 | 0.1812 | 0.0533 | **3.4003** | 1.5879 | 7.2814 |
| J81 | J81.0 | Acute pulmonary edema | 2462 | 0.1767 | 0.0524 | **3.3700** | 2.7995 | 4.0567 |
| G20 | G20. | Parkinson's disease | 5105 | 0.1732 | 0.0515 | **3.3623** | 2.9456 | 3.8379 |
| N25 | N25.81 | Secondary hyperparathyroidism of renal origin | 6475 | 0.1699 | 0.0511 | **3.3268** | 2.9515 | 3.7497 |
| J96 | J96.01 | Acute respiratory failure with hypoxia | 13350 | 0.1624 | 0.0489 | **3.3231** | 3.0408 | 3.6316 |
| I12 | I12.9 | Hypertensive chronic kidney disease w stg 1-4/unsp chr kdny | 26422 | 0.1498 | 0.0452 | **3.3156** | 3.0895 | 3.5582 |
| D02 | D02.20 | Carcinoma in situ of unspecified bronchus and lung | 2023 | 0.1740 | 0.0526 | **3.3075** | 2.6919 | 4.0639 |
| W07 | W07.XX<br>XA | Fall from chair, initial encounter | 1948 | 0.1740 | 0.0526 | **3.3063** | 2.6807 | 4.0780 |
| C94 | C94.6 | Myelodysplastic disease, not classified | 412 | 0.1748 | 0.0532 | **3.2860** | 2.0929 | 5.1592 |
| N19 | N19. | Unspecified kidney failure | 7383 | 0.1663 | 0.0508 | **3.2729** | 2.9203 | 3.6680 |
| L12 | L12.0 | Bullous pemphigoid | 334 | 0.1737 | 0.0532 | **3.2634** | 1.9740 | 5.3952 |
| R26 | R26.9 | Unspecified abnormalities of gait and mobility | 67067 | 0.1202 | 0.0369 | **3.2578** | 3.0675 | 3.4598 |
| Z49 | Z49.31 | Encounter for adequacy testing for hemodialysis | 1540 | 0.1714 | 0.0528 | **3.2472** | 2.5607 | 4.1178 |
| I96 | I96. | Gangrene, not elsewhere classified | 2079 | 0.1708 | 0.0526 | **3.2459** | 2.6429 | 3.9864 |
| H27 | H27.8 | Other specified disorders of lens | 6825 | 0.1642 | 0.0511 | **3.2170** | 2.8561 | 3.6236 |
| I25 | I25.10 | Atherosclerotic heart disease of native coronary artery w/o ang pctrs | 77259 | 0.1139 | 0.0355 | **3.2083** | 3.0212 | 3.4069 |
| T66 | T66.XXX<br>A | Radiation sickness, unspecified, initial encounter | 211 | 0.1706 | 0.0533 | **3.2036** | 1.6912 | 6.0686 |
| C34 | C34.90 | Malignant neoplasm of unsp part of unsp bronchus or lung | 3805 | 0.1664 | 0.0520 | **3.1962** | 2.7354 | 3.7346 |
| E46 | E46. | Unspecified protein-calorie malnutrition | 3568 | 0.1659 | 0.0521 | **3.1825** | 2.7098 | 3.7377 |
| E44 | E44.0 | Moderate protein-calorie malnutrition | 3752 | 0.1647 | 0.0521 | **3.1623** | 2.7011 | 3.7022 |
| C91 | C91.10 | Chronic lymphocytic leuk of B-cell type not achieve remis | 1854 | 0.1661 | 0.0527 | **3.1517** | 2.5270 | 3.9307 |
| N17 | N17.9 | Acute kidney failure, unspecified | 31785 | 0.1396 | 0.0444 | **3.1421** | 2.9347 | 3.3641 |
| Z19 | Z19.2 | Hormone resistant malignancy status | 174 | 0.1667 | 0.0533 | **3.1286** | 1.5331 | 6.3847 |
| L22 | L22. | Diaper dermatitis | 774 | 0.1654 | 0.0531 | **3.1159** | 2.2159 | 4.3814 |
| G91 | G91.2 | (Idiopathic) normal pressure hydrocephalus | 917 | 0.1647 | 0.0530 | **3.1053** | 2.2680 | 4.2515 |
| G92 | G92. | Toxic encephalopathy | 1085 | 0.1631 | 0.0530 | **3.0793** | 2.3025 | 4.1180 |
| Y73 | Y73.2 | Prosth/oth implnt/mtrls gastroent and urol dev assoc w incdt | 735 | 0.1633 | 0.0531 | **3.0752** | 2.1619 | 4.3743 |
| C79 | C79.51 | Secondary malignant neoplasm of bone | 2724 | 0.1612 | 0.0525 | **3.0721** | 2.5499 | 3.7014 |
| Z96 | Z96.1 | Presence of intraocular lens | 58557 | 0.1206 | 0.0393 | **3.0656** | 2.8838 | 3.2588 |
| E78 | E78.5 | Hyperlipidemia, unspecified | 223873 | 0.0692 | 0.0226 | **3.0600** | 2.8037 | 3.3397 |
| I73 | I73.9 | Peripheral vascular disease, unspecified | 31350 | 0.1369 | 0.0448 | **3.0545** | 2.8506 | 3.2730 |
| R18 | R18.8 | Other ascites | 2408 | 0.1586 | 0.0526 | **3.0172** | 2.4710 | 3.6840 |
| C66 | C66.9 | Malignant neoplasm of unspecified ureter | 237 | 0.1603 | 0.0533 | **3.0107** | 1.6104 | 5.6286 |
| Z95 | Z95.1 | Presence of aortocoronary bypass graft | 34407 | 0.1330 | 0.0444 | **2.9986** | 2.8020 | 3.2089 |
| Z71 | Z71.89 | Other specified counseling | 308757 | 0.0568 | 0.0190 | **2.9898** | 2.5115 | 3.5592 |
| J91 | J91.8 | Pleural effusion in other conditions classified elsewhere | 4873 | 0.1545 | 0.0519 | **2.9798** | 2.5795 | 3.4422 |
| G11 | G11.1 | Early-onset cerebellar ataxia | 586 | 0.1570 | 0.0532 | **2.9538** | 1.9729 | 4.4224 |
| I48 | I48.91 | Unspecified atrial fibrillation | 35951 | 0.1301 | 0.0442 | **2.9419** | 2.7502 | 3.1470 |
| J15 | J15.9 | Unspecified bacterial pneumonia | 9858 | 0.1482 | 0.0505 | **2.9350** | 2.6389 | 3.2644 |
| N14 | N14.0 | Analgesic nephropathy | 405 | 0.1556 | 0.0532 | **2.9235** | 1.7952 | 4.7611 |
| I67 | I67.89 | Other cerebrovascular disease | 16692 | 0.1420 | 0.0487 | **2.9125** | 2.6714 | 3.1754 |
| H26 | H26.9 | Unspecified cataract | 60786 | 0.1156 | 0.0398 | **2.9053** | 2.7326 | 3.0889 |
| Y71 | Y71.2 | Prosth/oth implnt/mtrls cardiovascular devices assoc w incdt | 989 | 0.1537 | 0.0530 | **2.8978** | 2.1144 | 3.9715 |
| J17 | J17. | Pneumonia in diseases classified elsewhere | 2185 | 0.1524 | 0.0527 | **2.8925** | 2.3345 | 3.5838 |
| J61 | J61. | Pneumoconiosis due to asbestos and other mineral fibers | 1189 | 0.1531 | 0.0530 | **2.8892** | 2.1653 | 3.8551 |
| T82 | T82.818 A | Embolism due to vascular prosth dev/grft, initial encounter | 6593 | 0.1479 | 0.0515 | **2.8738** | 2.5282 | 3.2668 |
| I95 | I95.9 | Hypotension, unspecified | 32131 | 0.1301 | 0.0453 | **2.8710** | 2.6771 | 3.0789 |
| I70 | I70.0 | Atherosclerosis of aorta | 19489 | 0.1379 | 0.0482 | **2.8613** | 2.6354 | 3.1065 |
| I27 | I27.20 | Pulmonary hypertension, unspecified | 9999 | 0.1445 | 0.0506 | **2.8580** | 2.5677 | 3.1811 |
| W90 | W90.8XX A | Exposure to other nonionizing radiation, initial encounter | 250 | 0.1520 | 0.0533 | **2.8541** | 1.5219 | 5.3522 |
| I62 | I62.00 | Nontraumatic subdural hemorrhage, unspecified | 1571 | 0.1509 | 0.0529 | **2.8530** | 2.2142 | 3.6762 |
| N40 | N40.0 | Benign prostatic hyperplasia without lower urinry tract symp | 85767 | 0.1036 | 0.0363 | **2.8518** | 2.6852 | 3.0289 |
| R65 | R65.20 | Severe sepsis without septic shock | 5450 | 0.1468 | 0.0518 | **2.8334** | 2.4613 | 3.2617 |
| A41 | A41.9 | Sepsis, unspecified organism | 12759 | 0.1412 | 0.0499 | **2.8303** | 2.5681 | 3.1193 |
| C83 | C83.30 | Diffuse large B-cell lymphoma, unspecified site | 1195 | 0.1498 | 0.0530 | **2.8268** | 2.1123 | 3.7829 |
| L97 | L97.509 | Non-pressure chronic ulcer oth prt unsp foot w unsp severity | 13950 | 0.1401 | 0.0496 | **2.8231** | 2.5701 | 3.1010 |
| I06 | I06.0 | Rheumatic aortic stenosis | 614 | 0.1498 | 0.0532 | **2.8189** | 1.8796 | 4.2275 |
| C92 | C92.10 | Chronic myeloid leuk, BCR/ABL-positive, not achieve remis | 602 | 0.1495 | 0.0532 | **2.8123** | 1.8667 | 4.2369 |
| G31 | G31.84 | Mild cognitive impairment, so stated | 21486 | 0.1343 | 0.0479 | **2.8064** | 2.5905 | 3.0402 |
| G70 | G70.00 | Myasthenia gravis without (acute) exacerbation | 745 | 0.1490 | 0.0531 | **2.8049** | 1.9386 | 4.0581 |
| G21 | G21.11 | Neuroleptic induced parkinsonism | 2043 | 0.1478 | 0.0528 | **2.8019** | 2.2364 | 3.5103 |
| D61 | D61.818 | Other pancytopenia | 3153 | 0.1468 | 0.0525 | **2.7995** | 2.3311 | 3.3619 |
| J70 | J70.5 | Respiratory conditions due to smoke inhalation | 438 | 0.1484 | 0.0532 | **2.7891** | 1.7221 | 4.5172 |
| R57 | R57.0 | Cardiogenic shock | 1089 | 0.1478 | 0.0530 | **2.7881** | 2.0503 | 3.7914 |
| I63 | I63.9 | Cerebral infarction, unspecified | 18075 | 0.1357 | 0.0487 | **2.7853** | 2.5572 | 3.0338 |
| C19 | C19. | Malignant neoplasm of rectosigmoid junction | 547 | 0.1481 | 0.0532 | **2.7847** | 1.8074 | 4.2903 |
| H25 | H25.13 | Age-related nuclear cataract, bilateral | 127537 | 0.0888 | 0.0320 | **2.7742** | 2.6054 | 2.9539 |
| W08 | W08.XX XA | Fall from other furniture, initial encounter | 544 | 0.1471 | 0.0532 | **2.7653** | 1.7896 | 4.2730 |
| Z75 | Z75.1 | Person awaiting admission to adequate facility elsewhere | 10861 | 0.1394 | 0.0505 | **2.7610** | 2.4854 | 3.0672 |
| S88 | S88.119 A | Complete traum amp at lev betw kn & ankl, unsp low leg, init | 1412 | 0.1459 | 0.0529 | **2.7556** | 2.0985 | 3.6185 |
| J84 | J84.10 | Pulmonary fibrosis, unspecified | 7398 | 0.1414 | 0.0514 | **2.7524** | 2.4303 | 3.1172 |
| N08 | N08. | Glomerular disorders in diseases classified elsewhere | 2453 | 0.1443 | 0.0527 | **2.7400** | 2.2233 | 3.3768 |
| I65 | I65.29 | Occlusion and stenosis of unspecified carotid artery | 16848 | 0.1344 | 0.0491 | **2.7367** | 2.5052 | 2.9897 |
| S78 | S78.019 A | Complete traumatic amputation at unsp hip joint, init encntr | 447 | 0.1454 | 0.0532 | **2.7329** | 1.6860 | 4.4299 |
| J92 | J92.9 | Pleural plaque without asbestos | 1007 | 0.1450 | 0.0531 | **2.7326** | 1.9780 | 3.7751 |
| I69 | I69.898 | Other sequelae of other cerebrovascular disease | 14535 | 0.1356 | 0.0497 | **2.7310** | 2.4866 | 2.9995 |
| D64 | D64.9 | Anemia, unspecified | 62258 | 0.1103 | 0.0405 | **2.7201** | 2.5576 | 2.8930 |
| S72 | S72.009<br>A | Fracture of unsp part of neck of<br>unsp femur, init | 3337 | 0.1423 | 0.0524 | **2.7140** | 2.2641 | 3.2535 |
| C80 | C80.1 | Malignant (primary) neoplasm,<br>unspecified | 1345 | 0.1435 | 0.0530 | **2.7089** | 2.0438 | 3.5904 |
| W05 | W05.0XX<br>A | Fall from non-moving wheelchair,<br>initial encounter | 660 | 0.1439 | 0.0532 | **2.7080** | 1.8148 | 4.0408 |
| S98 | S98.119<br>A | Complete traumatic amputation<br>of unsp great toe, init encntr | 1353 | 0.1434 | 0.0530 | **2.7069** | 2.0437 | 3.5853 |
| R33 | R33.9 | Retention of urine, unspecified | 20323 | 0.1307 | 0.0484 | **2.7009** | 2.4861 | 2.9343 |
| C71 | C71.9 | Malignant neoplasm of brain,<br>unspecified | 557 | 0.1436 | 0.0532 | **2.7007** | 1.7462 | 4.1768 |
| J47 | J47.9 | Bronchiectasis, uncomplicated | 1749 | 0.1424 | 0.0529 | **2.6928** | 2.1001 | 3.4529 |
| D53 | D53.9 | Nutritional anemia, unspecified | 4238 | 0.1406 | 0.0522 | **2.6927** | 2.2886 | 3.1682 |
| J44 | J44.9 | Chronic obstructive pulmonary<br>disease, unspecified | 63653 | 0.1089 | 0.0405 | **2.6858** | 2.5255 | 2.8563 |
| I79 | I79.8 | Oth disord of art,arterioles &<br>capilare in dis classd elswhr | 940 | 0.1426 | 0.0531 | **2.6855** | 1.9159 | 3.7643 |
| M80 | M80.88X<br>S | Oth osteopor w current path<br>fracture, vertebra(e), sequela | 1227 | 0.1418 | 0.0530 | **2.6752** | 1.9881 | 3.5997 |
| I44 | I44.0 | Atrioventricular block, first<br>degree | 14983 | 0.1324 | 0.0497 | **2.6653** | 2.4270 | 2.9270 |
| N26 | N26.1 | Atrophy of kidney (terminal) | 757 | 0.1413 | 0.0531 | **2.6602** | 1.8232 | 3.8816 |
| J80 | J80. | Acute respiratory distress<br>syndrome | 1161 | 0.1404 | 0.0530 | **2.6474** | 1.9479 | 3.5981 |
| C90 | C90.00 | Multiple myeloma not having<br>achieved remission | 1041 | 0.1402 | 0.0531 | **2.6431** | 1.9115 | 3.6547 |
| C22 | C22.0 | Liver cell carcinoma | 1150 | 0.1400 | 0.0530 | **2.6397** | 1.9385 | 3.5947 |
| Y82 | Y82.8 | Other medical devices<br>associated with adverse<br>incidents | 1007 | 0.1400 | 0.0531 | **2.6383** | 1.8972 | 3.6688 |
| I35 | I35.0 | Nonrheumatic aortic (valve)<br>stenosis | 12454 | 0.1320 | 0.0503 | **2.6225** | 2.3688 | 2.9033 |
| C32 | C32.9 | Malignant neoplasm of larynx,<br>unspecified | 930 | 0.1387 | 0.0531 | **2.6125** | 1.8504 | 3.6883 |
| I08 | I08.0 | Rheumatic disorders of both<br>mitral and aortic valves | 3548 | 0.1370 | 0.0524 | **2.6117** | 2.1819 | 3.1263 |
| E11 | E11.9 | Type 2 diabetes mellitus without<br>complications | 114394 | 0.0902 | 0.0346 | **2.6085** | 2.4536 | 2.7731 |
| I22 | I22.2 | Subsequent non-ST elevation<br>(NSTEMI) myocardial infarction | 897 | 0.1382 | 0.0531 | **2.6031** | 1.8311 | 3.7006 |
| I71 | I71.4 | Abdominal aortic aneurysm,<br>without rupture | 12513 | 0.1311 | 0.0504 | **2.6027** | 2.3505 | 2.8818 |
| I21 | I21.4 | Non-ST elevation (NSTEMI)<br>myocardial infarction | 14174 | 0.1297 | 0.0500 | **2.5947** | 2.3550 | 2.8588 |
| N04 | N04.9 | Nephrotic syndrome with<br>unspecified morphologic<br>changes | 1098 | 0.1375 | 0.0531 | **2.5920** | 1.8837 | 3.5666 |
| J94 | J94.8 | Other specified pleural conditions | 726 | 0.1377 | 0.0531 | **2.5916** | 1.7519 | 3.8338 |
| D04 | D04.9 | Carcinoma in situ of skin,<br>unspecified | 3845 | 0.1355 | 0.0524 | **2.5864** | 2.1733 | 3.0780 |
| E87 | E87.6 | Hypokalemia | 54353 | 0.1097 | 0.0426 | **2.5770** | 2.4185 | 2.7459 |
| R60 | R60.0 | Localized edema | 61977 | 0.1065 | 0.0415 | **2.5673** | 2.4126 | 2.7320 |
| T45 | T45.1X5<br>A | Adverse effect of antineoplastic<br>and immunosup drugs, init | 2220 | 0.1351 | 0.0528 | **2.5598** | 2.0383 | 3.2146 |
| W01 | W01.0XX<br>A | Fall same lev from slip/trip w/o<br>strike against object, init | 9034 | 0.1311 | 0.0512 | **2.5594** | 2.2742 | 2.8805 |
| C85 | C85.80 | Oth types of non-Hodgkin<br>lymphoma, unspecified site | 2228 | 0.1351 | 0.0528 | **2.5592** | 2.0386 | 3.2127 |
| H34 | H34.219 | Partial retinal artery occlusion, unspecified eye | 7070 | 0.1321 | 0.0517 | **2.5575** | 2.2414 | 2.9181 |
| J18 | J18.9 | Pneumonia, unspecified organism | 32422 | 0.1187 | 0.0464 | **2.5551** | 2.3768 | 2.7467 |
| Z45 | Z45.2 | Encounter for adjustment and management of VAD | 11371 | 0.1290 | 0.0507 | **2.5442** | 2.2854 | 2.8322 |
| I74 | I74.3 | Embolism and thrombosis of arteries of the lower extremities | 3254 | 0.1337 | 0.0526 | **2.5438** | 2.1033 | 3.0764 |
| T86 | T86.10 | Unspecified complication of kidney transplant | 1392 | 0.1343 | 0.0530 | **2.5349** | 1.9012 | 3.3798 |
| E08 | E08.42 | Diabetes due to underlying condition w diabetic polyneurop | 20437 | 0.1237 | 0.0488 | **2.5346** | 2.3283 | 2.7591 |
| J93 | J93.83 | Other pneumothorax | 1123 | 0.1345 | 0.0531 | **2.5341** | 1.8406 | 3.4889 |
| A40 | A40.9 | Streptococcal sepsis, unspecified | 789 | 0.1343 | 0.0531 | **2.5281** | 1.7269 | 3.7010 |
| C67 | C67.9 | Malignant neoplasm of bladder, unspecified | 4135 | 0.1323 | 0.0524 | **2.5266** | 2.1310 | 2.9956 |
| C18 | C18.9 | Malignant neoplasm of colon, unspecified | 4296 | 0.1318 | 0.0523 | **2.5179** | 2.1295 | 2.9771 |
| T44 | T44.7X5A | Adverse effect of beta-adrenoreceptor antagonists, init | 831 | 0.1336 | 0.0531 | **2.5139** | 1.7318 | 3.6493 |
| R27 | R27.0 | Ataxia, unspecified | 5819 | 0.1306 | 0.0520 | **2.5125** | 2.1722 | 2.9061 |
| Z93 | Z93.3 | Colostomy status | 4186 | 0.1312 | 0.0524 | **2.5048** | 2.1131 | 2.9692 |
| Z89 | Z89.519 | Acquired absence of unspecified leg below knee | 5730 | 0.1302 | 0.0520 | **2.5031** | 2.1612 | 2.8992 |
| Z43 | Z43.3 | Encounter for attention to colostomy | 2992 | 0.1317 | 0.0526 | **2.5019** | 2.0489 | 3.0550 |
| E10 | E10.9 | Type 1 diabetes mellitus without complications | 20517 | 0.1220 | 0.0489 | **2.4951** | 2.2911 | 2.7172 |
| H35 | H35.31 | Nonexudative age-related macular degeneration | 60393 | 0.1051 | 0.0421 | **2.4933** | 2.3418 | 2.6547 |
| I38 | I38. | Endocarditis, valve unspecified | 2607 | 0.1312 | 0.0527 | **2.4880** | 2.0084 | 3.0820 |
| I24 | I24.8 | Other forms of acute ischemic heart disease | 3221 | 0.1307 | 0.0526 | **2.4854** | 2.0481 | 3.0160 |
| C25 | C25.9 | Malignant neoplasm of pancreas, unspecified | 636 | 0.1321 | 0.0532 | **2.4835** | 1.6181 | 3.8117 |
| I77 | I77.1 | Stricture of artery | 7515 | 0.1280 | 0.0516 | **2.4789** | 2.1759 | 2.8241 |
| R32 | R32. | Unspecified urinary incontinence | 18819 | 0.1220 | 0.0493 | **2.4746** | 2.2651 | 2.7034 |
| I66 | I66.9 | Occlusion and stenosis of unspecified cerebral artery | 883 | 0.1314 | 0.0531 | **2.4728** | 1.7165 | 3.5623 |
| G12 | G12.21 | Amyotrophic lateral sclerosis | 685 | 0.1314 | 0.0532 | **2.4710** | 1.6331 | 3.7387 |
| I42 | I42.9 | Cardiomyopathy, unspecified | 15337 | 0.1234 | 0.0500 | **2.4677** | 2.2418 | 2.7164 |
| I72 | I72.3 | Aneurysm of iliac artery | 3524 | 0.1294 | 0.0525 | **2.4632** | 2.0446 | 2.9675 |
| I05 | I05.8 | Other rheumatic mitral valve diseases | 1041 | 0.1306 | 0.0531 | **2.4607** | 1.7558 | 3.4485 |
| F09 | F09. | Unsp mental disorder due to known physiological condition | 2150 | 0.1298 | 0.0528 | **2.4557** | 1.9380 | 3.1118 |
| R77 | R77.0 | Abnormality of albumin | 1284 | 0.1301 | 0.0530 | **2.4522** | 1.8076 | 3.3266 |
| D62 | D62. | Acute posthemorrhagic anemia | 6406 | 0.1266 | 0.0519 | **2.4383** | 2.1167 | 2.8087 |
| M86 | M86.9 | Osteomyelitis, unspecified | 7523 | 0.1252 | 0.0517 | **2.4219** | 2.1227 | 2.7632 |
| J43 | J43.9 | Emphysema, unspecified | 9950 | 0.1238 | 0.0512 | **2.4182** | 2.1523 | 2.7169 |
| I87 | I87.2 | Venous insufficiency (chronic) (peripheral) | 23285 | 0.1175 | 0.0486 | **2.4162** | 2.2250 | 2.6239 |
| M81 | M81.0 | Age-related osteoporosis w/o current pathological fracture | 10016 | 0.1236 | 0.0512 | **2.4143** | 2.1494 | 2.7118 |
| K72 | K72.90 | Hepatic failure, unspecified without coma | 1867 | 0.1275 | 0.0529 | **2.4089** | 1.8641 | 3.1128 |
| W19 | W19.XX<br>XA | Unspecified fall, initial encounter | 21689 | 0.1178 | 0.0489 | **2.4085** | 2.2128 | 2.6214 |
| D38 | D38.1 | Neoplasm of uncertain behavior of trachea, bronchus and lung | 1841 | 0.1271 | 0.0529 | **2.4015** | 1.8543 | 3.1101 |
| T83 | T83.098<br>A | Mech compl of other urinary catheter, initial encounter | 3957 | 0.1259 | 0.0525 | **2.3983** | 2.0056 | 2.8679 |
| I07 | I07.1 | Rheumatic tricuspid insufficiency | 3276 | 0.1258 | 0.0526 | **2.3898** | 1.9643 | 2.9075 |
| M31 | M31.6 | Other giant cell arteritis | 1113 | 0.1267 | 0.0531 | **2.3863** | 1.7117 | 3.3268 |
| T38 | T38.0X5<br>A | Adverse effect of glucocort/synth analog, init | 2095 | 0.1260 | 0.0529 | **2.3831** | 1.8674 | 3.0411 |
| E09 | E09.65 | Drug or chemical induced diabetes mellitus w hyperglycemia | 893 | 0.1265 | 0.0531 | **2.3814** | 1.6435 | 3.4506 |
| E63 | E63.9 | Nutritional deficiency, unspecified | 1355 | 0.1255 | 0.0530 | **2.3654** | 1.7469 | 3.2027 |
| H90 | H90.3 | Sensorineural hearing loss, bilateral | 118138 | 0.0855 | 0.0362 | **2.3616** | 2.2218 | 2.5102 |
| I61 | I61.9 | Nontraumatic intracerebral hemorrhage, unspecified | 1197 | 0.1253 | 0.0531 | **2.3610** | 1.7102 | 3.2595 |
| G46 | G46.4 | Cerebellar stroke syndrome | 4838 | 0.1234 | 0.0523 | **2.3586** | 2.0018 | 2.7791 |
| R13 | R13.10 | Dysphagia, unspecified | 40814 | 0.1080 | 0.0459 | **2.3554** | 2.1977 | 2.5245 |
| C20 | C20. | Malignant neoplasm of rectum | 935 | 0.1251 | 0.0531 | **2.3552** | 1.6352 | 3.3920 |
| E21 | E21.3 | Hyperparathyroidism, unspecified | 4412 | 0.1233 | 0.0524 | **2.3526** | 1.9817 | 2.7930 |
| K94 | K94.23 | Gastrostomy malfunction | 1165 | 0.1245 | 0.0531 | **2.3446** | 1.6888 | 3.2551 |
| S37 | S37.009<br>A | Unspecified injury of unspecified kidney, initial encounter | 989 | 0.1244 | 0.0531 | **2.3412** | 1.6399 | 3.3423 |
| S81 | S81.009<br>A | Unspecified open wound, unspecified knee, initial encounter | 8362 | 0.1208 | 0.0516 | **2.3395** | 2.0586 | 2.6588 |
| D89 | D89.9 | Disorder involving the immune mechanism, unspecified | 1590 | 0.1239 | 0.0530 | **2.3378** | 1.7633 | 3.0995 |
| G45 | G45.9 | Transient cerebral ischemic attack, unspecified | 14740 | 0.1178 | 0.0504 | **2.3365** | 2.1140 | 2.5823 |
| Y84 | Y84.8 | Oth medical procedures cause abn react/compl, w/o misadvnt | 3576 | 0.1228 | 0.0526 | **2.3343** | 1.9295 | 2.8240 |
| R47 | R47.1 | Dysarthria and anarthria | 8462 | 0.1204 | 0.0516 | **2.3330** | 2.0539 | 2.6500 |
| I45 | I45.10 | Unspecified right bundle-branch block | 15283 | 0.1173 | 0.0503 | **2.3302** | 2.1113 | 2.5718 |
| G81 | G81.90 | Hemiplegia, unspecified affecting unspecified side | 3445 | 0.1225 | 0.0526 | **2.3279** | 1.9171 | 2.8267 |
| R41 | R41.2 | Retrograde amnesia | 42168 | 0.1065 | 0.0458 | **2.3270** | 2.1720 | 2.4930 |
| I11 | I11.9 | Hypertensive heart disease without heart failure | 28332 | 0.1117 | 0.0480 | **2.3255** | 2.1510 | 2.5142 |
| Q84 | Q84.5 | Enlarged and hypertrophic nails | 1698 | 0.1231 | 0.0530 | **2.3233** | 1.7664 | 3.0557 |
| Q27 | Q27.9 | Congenital malformation of peripheral vascular system, unsp | 1138 | 0.1230 | 0.0531 | **2.3170** | 1.6589 | 3.2362 |
| J12 | J12.9 | Viral pneumonia, unspecified | 1835 | 0.1226 | 0.0530 | **2.3155** | 1.7777 | 3.0160 |
| T36 | T36.8X5<br>A | Adverse effect of other systemic antibiotics, init encntr | 1254 | 0.1228 | 0.0531 | **2.3139** | 1.6824 | 3.1826 |
| I99 | I99.8 | Other disorder of circulatory system | 4579 | 0.1212 | 0.0524 | **2.3130** | 1.9511 | 2.7419 |
| S32 | S32.009<br>A | Unsp fracture of unsp lumbar vertebra, init for clos fx | 4732 | 0.1211 | 0.0524 | **2.3121** | 1.9555 | 2.7338 |
| J95 | J95.811 | Postprocedural pneumothorax | 2669 | 0.1218 | 0.0528 | **2.3067** | 1.8499 | 2.8763 |
| D69 | D69.6 | Thrombocytopenia, unspecified | 15284 | 0.1161 | 0.0504 | **2.3056** | 2.0880 | 2.5458 |
| Z51 | Z51.89 | Encounter for other specified aftercare | 178296 | 0.0729 | 0.0317 | **2.3027** | 2.1516 | 2.4644 |
| Z99 | Z99.89 | Dependence on other enabling machines and devices | 21756 | 0.1133 | 0.0492 | **2.3027** | 2.1125 | 2.5100 |
| N05 | N05.9 | Unsp nephritic syndrome with unspecified morphologic changes | 1450 | 0.1221 | 0.0530 | **2.3017** | 1.7091 | 3.0996 |
| N03 | N03.9 | Chronic nephritic syndrome with unsp morphologic changes | 736 | 0.1223 | 0.0532 | **2.2994** | 1.5164 | 3.4866 |
| R34 | R34. | Anuria and oliguria | 720 | 0.1222 | 0.0532 | **2.2981** | 1.5085 | 3.5011 |
| D09 | D09.0 | Carcinoma in situ of bladder | 3148 | 0.1210 | 0.0527 | **2.2967** | 1.8724 | 2.8172 |
| C76 | C76.0 | Malignant neoplasm of head, face and neck | 3221 | 0.1208 | 0.0527 | **2.2923** | 1.8726 | 2.8061 |
| I37 | I37.0 | Nonrheumatic pulmonary valve stenosis | 1552 | 0.1211 | 0.0530 | **2.2847** | 1.7112 | 3.0505 |
| Z94 | Z94.0 | Kidney transplant status | 3033 | 0.1203 | 0.0527 | **2.2824** | 1.8526 | 2.8120 |
| J16 | J16.8 | Pneumonia due to other specified infectious organisms | 1240 | 0.1210 | 0.0531 | **2.2789** | 1.6494 | 3.1486 |
| A04 | A04.7 | Enterocolitis due to Clostridium difficile | 5175 | 0.1190 | 0.0523 | **2.2754** | 1.9351 | 2.6755 |
| Z85 | Z85.828 | Personal history of other malignant neoplasm of skin | 43622 | 0.1042 | 0.0458 | **2.2743** | 2.1233 | 2.4362 |
| L60 | L60.0 | Ingrowing nail | 53817 | 0.1007 | 0.0444 | **2.2688** | 2.1255 | 2.4219 |
| H59 | H59.032 | Cystoid macular edema following cataract surgery, left eye | 2323 | 0.1197 | 0.0529 | **2.2634** | 1.7830 | 2.8732 |
| C77 | C77.0 | Sec and unsp malig neoplasm of nodes of head, face and neck | 2569 | 0.1195 | 0.0528 | **2.2622** | 1.8024 | 2.8393 |
| R53 | R53.1 | Weakness | 65148 | 0.0971 | 0.0430 | **2.2599** | 2.1221 | 2.4067 |
| E86 | E86.0 | Dehydration | 28085 | 0.1091 | 0.0483 | **2.2594** | 2.0876 | 2.4453 |
| T17 | T17.300 A | Unsp foreign body in larynx causing asphyxiation, init | 842 | 0.1200 | 0.0532 | **2.2563** | 1.5221 | 3.3445 |
| R80 | R80.3 | Bence Jones proteinuria | 13651 | 0.1142 | 0.0508 | **2.2489** | 2.0245 | 2.4983 |
| Z79 | Z79.899 | Other long term (current) drug therapy | 147374 | 0.0777 | 0.0346 | **2.2438** | 2.1065 | 2.3899 |
| D47 | D47.2 | Monoclonal gammopathy | 5592 | 0.1166 | 0.0523 | **2.2306** | 1.9049 | 2.6119 |
| N31 | N31.9 | Neuromuscular dysfunction of bladder, unspecified | 7306 | 0.1158 | 0.0520 | **2.2287** | 1.9386 | 2.5621 |
| D50 | D50.9 | Iron deficiency anemia, unspecified | 31287 | 0.1065 | 0.0479 | **2.2208** | 2.0569 | 2.3978 |
| S51 | S51.809 A | Unspecified open wound of unspecified forearm, init encntr | 3177 | 0.1171 | 0.0527 | **2.2207** | 1.8053 | 2.7317 |
| R15 | R15.9 | Full incontinence of feces | 3962 | 0.1166 | 0.0526 | **2.2176** | 1.8405 | 2.6720 |
| I34 | I34.0 | Nonrheumatic mitral (valve) insufficiency | 11109 | 0.1137 | 0.0513 | **2.2167** | 1.9744 | 2.4887 |
| I89 | I89.0 | Lymphedema, not elsewhere classified | 5047 | 0.1161 | 0.0524 | **2.2165** | 1.8772 | 2.6172 |
| H54 | H54.7 | Unspecified visual loss | 25803 | 0.1080 | 0.0488 | **2.2108** | 2.0363 | 2.4001 |
| I15 | I15.8 | Other secondary hypertension | 4169 | 0.1161 | 0.0526 | **2.2092** | 1.8411 | 2.6510 |
| R58 | R58. | Hemorrhage, not elsewhere classified | 1357 | 0.1172 | 0.0531 | **2.2077** | 1.6115 | 3.0243 |
| D01 | D01.0 | Carcinoma in situ of colon | 2376 | 0.1166 | 0.0529 | **2.2045** | 1.7352 | 2.8007 |
| R63 | R63.4 | Abnormal weight loss | 34666 | 0.1046 | 0.0475 | **2.2026** | 2.0448 | 2.3726 |
| I36 | I36.1 | Nonrheumatic tricuspid (valve) insufficiency | 3914 | 0.1157 | 0.0526 | **2.2002** | 1.8226 | 2.6561 |
| J13 | J13. | Pneumonia due to Streptococcus pneumoniae | 1115 | 0.1166 | 0.0531 | **2.1948** | 1.5499 | 3.1081 |
| N28 | N28.9 | Disorder of kidney and ureter, unspecified | 27416 | 0.1067 | 0.0486 | **2.1941** | 2.0241 | 2.3783 |
| I43 | I43. | Cardiomyopathy in diseases classified elsewhere | 1050 | 0.1162 | 0.0531 | **2.1867** | 1.5269 | 3.1315 |
| D68 | D68.8 | Other specified coagulation defects | 5690 | 0.1142 | 0.0523 | **2.1845** | 1.8648 | 2.5591 |
| E83 | E83.42 | Hypomagnesemia | 22641 | 0.1079 | 0.0494 | **2.1827** | 2.0014 | 2.3806 |
| E16 | E16.2 | Hypoglycemia, unspecified | 9251 | 0.1126 | 0.0517 | **2.1799** | 1.9207 | 2.4742 |
| C44 | C44.91 | Basal cell carcinoma of skin, unspecified | 21770 | 0.1080 | 0.0496 | **2.1778** | 1.9940 | 2.3785 |
| H61 | H61.23 | Impacted cerumen, bilateral | 58221 | 0.0966 | 0.0444 | **2.1776** | 2.0411 | 2.3232 |
| I51 | I51.7 | Cardiomegaly | 24480 | 0.1066 | 0.0492 | **2.1674** | 1.9918 | 2.3584 |
| Z46 | Z46.1 | Encounter for fitting and adjustment of hearing aid | 98123 | 0.0863 | 0.0399 | **2.1625** | 2.0355 | 2.2975 |
| H52 | H52.4 | Presbyopia | 192666 | 0.0694 | 0.0323 | **2.1442** | 2.0003 | 2.2985 |
| D41 | D41.4 | Neoplasm of uncertain behavior of bladder | 3097 | 0.1130 | 0.0528 | **2.1412** | 1.7289 | 2.6517 |
| E20 | E20.1 | Pseudohypoparathyroidism | 1647 | 0.1135 | 0.0530 | **2.1408** | 1.6002 | 2.8640 |
| C61 | C61. | Malignant neoplasm of prostate | 19707 | 0.1069 | 0.0500 | **2.1359** | 1.9473 | 2.3427 |
| K55 | K55.20 | Angiodysplasia of colon without hemorrhage | 2499 | 0.1128 | 0.0529 | **2.1336** | 1.6821 | 2.7064 |
| B96 | B96.81 | Helicobacter pylori as the cause of diseases classd elswhr | 20262 | 0.1066 | 0.0500 | **2.1330** | 1.9466 | 2.3373 |
| B95 | B95.61 | Methicillin suscep staph infct causing dis classd elswhr | 13437 | 0.1086 | 0.0511 | **2.1267** | 1.9078 | 2.3708 |
| N32 | N32.81 | Overactive bladder | 19165 | 0.1063 | 0.0502 | **2.1188** | 1.9291 | 2.3272 |
| E13 | E13.9 | Other specified diabetes mellitus without complications | 11229 | 0.1089 | 0.0514 | **2.1177** | 1.8824 | 2.3824 |
| T85 | T85.398 A | Mech compl of ocular prosth dev/grft, init | 3710 | 0.1113 | 0.0527 | **2.1128** | 1.7342 | 2.5739 |
| I85 | I85.00 | Esophageal varices without bleeding | 2480 | 0.1117 | 0.0529 | **2.1114** | 1.6608 | 2.6842 |
| W18 | W18.49X A | Oth slipping, tripping and stumbling w/o falling, init | 13407 | 0.1077 | 0.0511 | **2.1079** | 1.8898 | 2.3511 |
| R29 | R29.6 | Repeated falls | 19639 | 0.1055 | 0.0501 | **2.1035** | 1.9164 | 2.3089 |
| H02 | H02.839 | Dermatochalasis of unspecified eye, unspecified eyelid | 29211 | 0.1024 | 0.0487 | **2.1019** | 1.9401 | 2.2772 |
| B35 | B35.1 | Tinea unguium | 95767 | 0.0854 | 0.0407 | **2.0963** | 1.9729 | 2.2274 |
| G99 | G99.0 | Autonomic neuropathy in diseases classified elsewhere | 4227 | 0.1100 | 0.0526 | **2.0908** | 1.7351 | 2.5194 |
| K26 | K26.9 | Duodenal ulcer, unsp as acute or chronic, w/o hemor or perf | 3428 | 0.1103 | 0.0527 | **2.0904** | 1.7008 | 2.5693 |
| I31 | I31.9 | Disease of pericardium, unspecified | 3328 | 0.1103 | 0.0528 | **2.0899** | 1.6953 | 2.5764 |
| H49 | H49.20 | Sixth [abducent] nerve palsy, unspecified eye | 1988 | 0.1107 | 0.0530 | **2.0883** | 1.5957 | 2.7330 |
| T87 | T87.89 | Other complications of amputation stump | 1707 | 0.1107 | 0.0530 | **2.0875** | 1.5620 | 2.7898 |
| L62 | L62. | Nail disorders in diseases classified elsewhere | 1530 | 0.1105 | 0.0531 | **2.0813** | 1.5318 | 2.8279 |
| N47 | N47.6 | Balanoposthitis | 6273 | 0.1086 | 0.0523 | **2.0761** | 1.7776 | 2.4247 |
| N39 | N39.0 | Urinary tract infection, site not specified | 64087 | 0.0920 | 0.0444 | **2.0733** | 1.9446 | 2.2105 |
| T46 | T46.4X5 A | Adverse effect of angiotens-convert-enzyme inhibitors, init | 1931 | 0.1098 | 0.0530 | **2.0712** | 1.5746 | 2.7244 |
| Z16 | Z16.11 | Resistance to penicillins | 2546 | 0.1092 | 0.0529 | **2.0638** | 1.6235 | 2.6234 |
| I82 | I82.409 | Acute embolism and thombos unsp deep vn unsp lower extremity | 14028 | 0.1046 | 0.0511 | **2.0456** | 1.8351 | 2.2803 |
| N12 | N12. | Tubulo-interstitial nephritis, not spcf as acute or chronic | 1965 | 0.1084 | 0.0530 | **2.0449** | 1.5552 | 2.6887 |
| S91 | S91.309 A | Unspecified open wound, unspecified foot, initial encounter | 11184 | 0.1054 | 0.0516 | **2.0447** | 1.8135 | 2.3054 |
| H21 | H21.81 | Floppy iris syndrome | 5158 | 0.1072 | 0.0525 | **2.0422** | 1.7199 | 2.4248 |
| H01 | H01.009 | Unspecified blepharitis unspecified eye, unspecified eyelid | 31384 | 0.0992 | 0.0487 | **2.0391** | 1.8843 | 2.2066 |
| I97 | I97.710 | Intraoperative cardiac arrest during cardiac surgery | 2682 | 0.1078 | 0.0529 | **2.0371** | 1.6095 | 2.5783 |
| L85 | L85.3 | Xerosis cutis | 31066 | 0.0991 | 0.0487 | **2.0350** | 1.8799 | 2.2029 |
| M15 | M15.9 | Polyosteoarthritis, unspecified | 70950 | 0.0892 | 0.0439 | **2.0333** | 1.9090 | 2.1656 |
| I20 | I20.8 | Other forms of angina pectoris | 31188 | 0.0990 | 0.0487 | **2.0318** | 1.8770 | 2.1993 |
| N13 | N13.30 | Unspecified hydronephrosis | 13082 | 0.1040 | 0.0513 | **2.0264** | 1.8109 | 2.2676 |
| L57 | L57.0 | Actinic keratosis | 46813 | 0.0945 | 0.0467 | **2.0227** | 1.8866 | 2.1685 |
| G60 | G60.9 | Hereditary and idiopathic neuropathy, unspecified | 30297 | 0.0988 | 0.0489 | **2.0219** | 1.8661 | 2.1907 |
| M10 | M10.9 | Gout, unspecified | 32778 | 0.0981 | 0.0485 | **2.0210** | 1.8693 | 2.1849 |
| M1A | M1A.9XX<br>0 | Chronic gout, unspecified, without tophus (tophi) | 5350 | 0.1060 | 0.0525 | **2.0192** | 1.7039 | 2.3928 |
| I26 | I26.99 | Other pulmonary embolism without acute cor pulmonale | 7323 | 0.1051 | 0.0522 | **2.0148** | 1.7400 | 2.3330 |
| K56 | K56.60 | Unspecified intestinal obstruction | 8535 | 0.1047 | 0.0520 | **2.0141** | 1.7569 | 2.3090 |
| G58 | G58.9 | Mononeuropathy, unspecified | 20738 | 0.1011 | 0.0502 | **2.0134** | 1.8348 | 2.2093 |
| D30 | D30.3 | Benign neoplasm of bladder | 1931 | 0.1067 | 0.0530 | **2.0119** | 1.5228 | 2.6581 |
| K59 | K59.00 | Constipation, unspecified | 59540 | 0.0911 | 0.0453 | **2.0108** | 1.8833 | 2.1469 |
| R01 | R01.1 | Cardiac murmur, unspecified | 14466 | 0.1028 | 0.0511 | **2.0104** | 1.8045 | 2.2398 |
| I47 | I47.1 | Supraventricular tachycardia | 12549 | 0.1031 | 0.0514 | **2.0053** | 1.7873 | 2.2500 |
| L11 | L11.0 | Acquired keratosis follicularis | 16439 | 0.1020 | 0.0509 | **2.0047** | 1.8095 | 2.2209 |
| I49 | I49.9 | Cardiac arrhythmia, unspecified | 59805 | 0.0907 | 0.0453 | **2.0015** | 1.8746 | 2.1370 |
| M62 | M62.81 | Muscle weakness (generalized) | 71400 | 0.0882 | 0.0441 | **2.0014** | 1.8789 | 2.1319 |
| G82 | G82.20 | Paraplegia, unspecified | 2430 | 0.1058 | 0.0530 | **1.9973** | 1.5557 | 2.5643 |
| J98 | J98.4 | Other disorders of lung | 35483 | 0.0964 | 0.0483 | **1.9959** | 1.8494 | 2.1540 |
| C64 | C64.9 | Malignant neoplasm of unsp kidney, except renal pelvis | 3149 | 0.1048 | 0.0528 | **1.9829** | 1.5896 | 2.4736 |
| H91 | H91.90 | Unspecified hearing loss, unspecified ear | 71156 | 0.0876 | 0.0443 | **1.9790** | 1.8576 | 2.1083 |
| E53 | E53.8 | Deficiency of other specified B group vitamins | 19291 | 0.0996 | 0.0505 | **1.9712** | 1.7899 | 2.1710 |
| R44 | R44.0 | Auditory hallucinations | 4298 | 0.1038 | 0.0527 | **1.9697** | 1.6273 | 2.3841 |
| R97 | R97.2 | Elevated prostate specific antigen [PSA] | 36803 | 0.0950 | 0.0483 | **1.9694** | 1.8259 | 2.1241 |
| R91 | R91.8 | Other nonspecific abnormal finding of lung field | 37585 | 0.0947 | 0.0482 | **1.9658** | 1.8235 | 2.1193 |
| H40 | H40.009 | Preglaucoma, unspecified, unspecified eye | 61759 | 0.0889 | 0.0454 | **1.9583** | 1.8344 | 2.0905 |
| I16 | I16.0 | Hypertensive urgency | 5096 | 0.1024 | 0.0526 | **1.9481** | 1.6320 | 2.3254 |
| Q61 | Q61.00 | Congenital renal cyst, unspecified | 7172 | 0.1018 | 0.0523 | **1.9467** | 1.6744 | 2.2634 |
| D49 | D49.2 | Neoplasm of unsp behavior of bone, soft tissue, and skin | 14001 | 0.0998 | 0.0513 | **1.9437** | 1.7390 | 2.1726 |
| Z76 | Z76.0 | Encounter for issue of repeat prescription | 161461 | 0.0715 | 0.0369 | **1.9405** | 1.8214 | 2.0674 |
| D07 | D07.5 | Carcinoma in situ of prostate | 9818 | 0.1006 | 0.0519 | **1.9381** | 1.7008 | 2.2086 |
| K74 | K74.60 | Unspecified cirrhosis of liver | 8439 | 0.1010 | 0.0521 | **1.9372** | 1.6840 | 2.2285 |
| K83 | K83.1 | Obstruction of bile duct | 3048 | 0.1024 | 0.0529 | **1.9355** | 1.5416 | 2.4301 |
| Z86 | Z86.010 | Personal history of colonic polyps | 103774 | 0.0803 | 0.0415 | **1.9350** | 1.8211 | 2.0561 |
| N42 | N42.9 | Disorder of prostate, unspecified | 10264 | 0.1003 | 0.0519 | **1.9328** | 1.7003 | 2.1972 |
| Y83 | Y83.1 | Implnt of artif int dev cause abn react/compl, w/o misadvnt | 11053 | 0.0999 | 0.0518 | **1.9296** | 1.7044 | 2.1845 |
| L84 | L84. | Corns and callosities | 31962 | 0.0946 | 0.0490 | **1.9284** | 1.7801 | 2.0891 |
| R04 | R04.0 | Epistaxis | 14831 | 0.0988 | 0.0513 | **1.9272** | 1.7282 | 2.1491 |
| D72 | D72.829 | Elevated white blood cell count, unspecified | 25660 | 0.0958 | 0.0499 | **1.9221** | 1.7619 | 2.0968 |
| G93 | G93.3 | Postviral fatigue syndrome | 39656 | 0.0925 | 0.0482 | **1.9201** | 1.7824 | 2.0685 |
| B02 | B02.9 | Zoster without complications | 16479 | 0.0976 | 0.0511 | **1.9118** | 1.7221 | 2.1225 |
| Z92 | Z92.3 | Personal history of irradiation | 8961 | 0.0994 | 0.0521 | **1.9092** | 1.6643 | 2.1900 |
| N30 | N30.00 | Acute cystitis without hematuria | 10151 | 0.0988 | 0.0519 | **1.9027** | 1.6710 | 2.1666 |
| R78 | R78.89 | Finding of oth substances, not normally found in blood | 32503 | 0.0933 | 0.0491 | **1.9012** | 1.7551 | 2.0595 |
| R55 | R55. | Syncope and collapse | 30812 | 0.0937 | 0.0493 | **1.9004** | 1.7516 | 2.0619 |
| E03 | E03.9 | Hypothyroidism, unspecified | 35891 | 0.0925 | 0.0487 | **1.9000** | 1.7588 | 2.0526 |
| H04 | H04.123 | Dry eye syndrome of bilateral lacrimal glands | 82128 | 0.0830 | 0.0439 | **1.8911** | 1.7770 | 2.0126 |
| K27 | K27.9 | Peptic ulc, site unsp, unsp as ac or chr, w/o hemor or perf | 7919 | 0.0987 | 0.0522 | **1.8901** | 1.6332 | 2.1874 |
| Z60 | Z60.2 | Problems related to living alone | 11811 | 0.0974 | 0.0517 | **1.8817** | 1.6657 | 2.1257 |
| E27 | E27.9 | Disorder of adrenal gland, unspecified | 3916 | 0.0991 | 0.0528 | **1.8767** | 1.5290 | 2.3034 |
| F06 | F06.30 | Mood disorder due to known physiological condition, unsp | 31408 | 0.0926 | 0.0493 | **1.8761** | 1.7295 | 2.0351 |
| Z65 | Z65.9 | Problem related to unspecified psychosocial circumstances | 127694 | 0.0753 | 0.0401 | **1.8759** | 1.7653 | 1.9935 |
| K25 | K25.9 | Gastric ulcer, unsp as acute or chronic, w/o hemor or perf | 5442 | 0.0985 | 0.0526 | **1.8727** | 1.5718 | 2.2312 |
| K86 | K86.1 | Other chronic pancreatitis | 5781 | 0.0983 | 0.0526 | **1.8696** | 1.5768 | 2.2168 |
| K68 | K68.11 | Postprocedural retroperitoneal abscess | 4165 | 0.0984 | 0.0528 | **1.8654** | 1.5280 | 2.2774 |
| H18 | H18.51 | Endothelial corneal dystrophy | 11506 | 0.0961 | 0.0518 | **1.8546** | 1.6380 | 2.0999 |
| T81 | T81.89X<br>A | Oth complications of procedures, NEC, init | 10309 | 0.0957 | 0.0520 | **1.8411** | 1.6150 | 2.0989 |
| D51 | D51.9 | Vitamin B12 deficiency anemia, unspecified | 14069 | 0.0948 | 0.0515 | **1.8398** | 1.6416 | 2.0619 |
| D29 | D29.1 | Benign neoplasm of prostate | 6819 | 0.0963 | 0.0524 | **1.8370** | 1.5670 | 2.1535 |
| Z44 | Z44.8 | Encounter for fit/adjust of external prosthetic devices | 5211 | 0.0967 | 0.0527 | **1.8369** | 1.5333 | 2.2005 |
| M84 | M84.369<br>S | Stress fracture, unspecified tibia and fibula, sequela | 5832 | 0.0965 | 0.0526 | **1.8362** | 1.5472 | 2.1791 |
| C43 | C43.9 | Malignant melanoma of skin, unspecified | 4224 | 0.0968 | 0.0528 | **1.8345** | 1.5020 | 2.2405 |
| R82 | R82.5 | Elevated urine levels of drug/meds/biol subst | 14186 | 0.0945 | 0.0515 | **1.8328** | 1.6357 | 2.0537 |
| R35 | R35.0 | Frequency of micturition | 39834 | 0.0890 | 0.0486 | **1.8321** | 1.6989 | 1.9756 |
| L82 | L82.1 | Other seborrheic keratosis | 55603 | 0.0859 | 0.0470 | **1.8281** | 1.7077 | 1.9571 |
| Z91 | Z91.19 | Patient's noncompliance w oth medical treatment and regimen | 64332 | 0.0842 | 0.0461 | **1.8244** | 1.7083 | 1.9482 |
| R49 | R49.0 | Dysphonia | 12308 | 0.0942 | 0.0518 | **1.8197** | 1.6114 | 2.0550 |
| G62 | G62.9 | Polyneuropathy, unspecified | 14108 | 0.0937 | 0.0516 | **1.8167** | 1.6201 | 2.0371 |
| H43 | H43.819 | Vitreous degeneration, unspecified eye | 38102 | 0.0887 | 0.0489 | **1.8155** | 1.6812 | 1.9606 |
| I83 | I83.90 | Asymptomatic varicose veins of unspecified lower extremity | 14263 | 0.0934 | 0.0516 | **1.8107** | 1.6154 | 2.0297 |
| S22 | S22.39X<br>A | Fracture of one rib, unsp side, init for clos fx | 10436 | 0.0942 | 0.0520 | **1.8102** | 1.5873 | 2.0644 |
| S31 | S31.000<br>A | Unsp opn wnd low back and pelv w/o penet retroperiton, init | 10430 | 0.0942 | 0.0520 | **1.8093** | 1.5864 | 2.0635 |
| Z13 | Z13.6 | Encounter for screening for cardiovascular disorders | 237211 | 0.0616 | 0.0341 | **1.8054** | 1.6707 | 1.9509 |
| K57 | K57.30 | Dvrtclos of lg int w/o perforation or abscess w/o bleeding | 60553 | 0.0842 | 0.0466 | **1.8047** | 1.6880 | 1.9295 |
| R06 | R06.02 | Shortness of breath | 118999 | 0.0750 | 0.0417 | **1.7984** | 1.6926 | 1.9108 |
| R09 | R09.81 | Nasal congestion | 40481 | 0.0871 | 0.0488 | **1.7869** | 1.6567 | 1.9274 |
| H31 | H31.009 | Unspecified chorioretinal scars, unspecified eye | 9734 | 0.0931 | 0.0522 | **1.7845** | 1.5566 | 2.0458 |
| Z23 | Z23. | Encounter for immunization | 260232 | 0.0594 | 0.0336 | **1.7694** | 1.6229 | 1.9292 |
| D12 | D12.0 | Benign neoplasm of cecum | 65120 | 0.0821 | 0.0465 | **1.7646** | 1.6519 | 1.8850 |
| K92 | K92.1 | Melena | 40170 | 0.0860 | 0.0489 | **1.7578** | 1.6287 | 1.8971 |
| S01 | S01.00X<br>A | Unspecified open wound of scalp, initial encounter | 7728 | 0.0921 | 0.0524 | **1.7574** | 1.5076 | 2.0485 |
| L21 | L21.9 | Seborrheic dermatitis, unspecified | 25335 | 0.0885 | 0.0505 | **1.7534** | 1.6012 | 1.9200 |
| R79 | R79.89 | Other specified abnormal findings of blood chemistry | 28403 | 0.0879 | 0.0502 | **1.7521** | 1.6065 | 1.9110 |
| K80 | K80.20 | Calculus of gallbladder w/o cholecystitis w/o obstruction | 16844 | 0.0896 | 0.0514 | **1.7429** | 1.5645 | 1.9417 |
| R31 | R31.9 | Hematuria, unspecified | 42562 | 0.0844 | 0.0489 | **1.7277** | 1.6025 | 1.8627 |
| L03 | L03.90 | Cellulitis, unspecified | 70833 | 0.0800 | 0.0463 | **1.7269** | 1.6183 | 1.8429 |
| R42 | R42. | Dizziness and giddiness | 67634 | 0.0800 | 0.0467 | **1.7131** | 1.6040 | 1.8297 |
| D48 | D48.5 | Neoplasm of uncertain behavior of skin | 44749 | 0.0834 | 0.0488 | **1.7106** | 1.5883 | 1.8424 |
| Z98 | Z98.890 | Other specified postprocedural states | 92911 | 0.0763 | 0.0447 | **1.7083** | 1.6057 | 1.8174 |
| R39 | R39.15 | Urgency of urination | 29413 | 0.0858 | 0.0503 | **1.7069** | 1.5654 | 1.8611 |
| K31 | K31.89 | Other diseases of stomach and duodenum | 15241 | 0.0872 | 0.0517 | **1.6854** | 1.5028 | 1.8900 |
| M89 | M89.9 | Disorder of bone, unspecified | 17841 | 0.0863 | 0.0515 | **1.6760** | 1.5056 | 1.8657 |
| R25 | R25.2 | Cramp and spasm | 28206 | 0.0846 | 0.0505 | **1.6743** | 1.5323 | 1.8294 |
| R69 | R69. | Illness, unspecified | 161563 | 0.0675 | 0.0405 | **1.6691** | 1.5685 | 1.7761 |
| Z09 | Z09. | Encntr for f/u exam aft trtmt for cond oth than malig neopl | 55214 | 0.0803 | 0.0481 | **1.6682** | 1.5555 | 1.7891 |
| Z48 | Z48.02 | Encounter for removal of sutures | 78659 | 0.0770 | 0.0462 | **1.6660** | 1.5627 | 1.7760 |
| M20 | M20.40 | Other hammer toe(s) (acquired), unspecified foot | 43692 | 0.0811 | 0.0492 | **1.6482** | 1.5280 | 1.7779 |
| L98 | L98.9 | Disorder of the skin and subcutaneous tissue, unspecified | 55257 | 0.0784 | 0.0485 | **1.6182** | 1.5080 | 1.7365 |
| M19 | M19.90 | Unspecified osteoarthritis, unspecified site | 89625 | 0.0737 | 0.0460 | **1.6021** | 1.5046 | 1.7059 |
| F12 | F12.10 | Cannabis abuse, uncomplicated | 24113 | 0.0346 | 0.0548 | **0.6324** | 0.5463 | 0.7320 |
| K58 | K58.9 | Irritable bowel syndrome without diarrhea | 17352 | 0.0329 | 0.0544 | **0.6046** | 0.5071 | 0.7207 |
| R51 | R51. | Headache | 51370 | 0.0335 | 0.0569 | **0.5896** | 0.5310 | 0.6546 |
| L70 | L70.0 | Acne vulgaris | 12335 | 0.0317 | 0.0541 | **0.5854** | 0.4739 | 0.7233 |
| M22 | M22.40 | Chondromalacia patellae, unspecified knee | 16168 | 0.0317 | 0.0544 | **0.5820** | 0.4835 | 0.7005 |
| F15 | F15.20 | Other stimulant dependence, uncomplicated | 8910 | 0.0300 | 0.0540 | **0.5554** | 0.4302 | 0.7168 |
| Z56 | Z56.0 | Unemployment, unspecified | 34996 | 0.0298 | 0.0560 | **0.5319** | 0.4661 | 0.6069 |
| A63 | A63.0 | Anogenital (venereal) warts | 4871 | 0.0279 | 0.0537 | **0.5199** | 0.3639 | 0.7428 |
| T74 | T74.21X<br>A | Adult sexual abuse, confirmed, initial encounter | 3385 | 0.0278 | 0.0536 | **0.5182** | 0.3376 | 0.7955 |
| Z20 | Z20.828 | Contact w and exposure to oth viral communicable diseases | 19317 | 0.0279 | 0.0549 | **0.5086** | 0.4243 | 0.6096 |
| X50 | X50.0XX<br>A | Overexertion from strenuous movement or load, init | 3150 | 0.0244 | 0.0536 | **0.4561** | 0.2838 | 0.7327 |
| J03 | J03.90 | Acute tonsillitis, unspecified | 5855 | 0.0243 | 0.0538 | **0.4505** | 0.3175 | 0.6391 |
| A60 | A60.9 | Anogenital herpesviral infection, unspecified | 7165 | 0.0241 | 0.0540 | **0.4475** | 0.3258 | 0.6145 |
| Z31 | Z31.5 | Encounter for procreative genetic counseling | 2984 | 0.0221 | 0.0536 | **0.4126** | 0.2471 | 0.6889 |
| G43 | G43.909 | Migraine, unsp, not intractable, without status migrainosus | 42103 | 0.0231 | 0.0576 | **0.4016** | 0.3503 | 0.4605 |
| N95 | N95.1 | Menopausal and female climacteric states | 9036 | 0.0216 | 0.0542 | **0.3982** | 0.2952 | 0.5371 |
| R92 | R92.8 | Oth abn and inconclusive findings on dx imaging of breast | 3749 | 0.0195 | 0.0537 | **0.3626** | 0.2225 | 0.5907 |
| N60 | N60.19 | Diffuse cystic mastopathy of unspecified breast | 3693 | 0.0187 | 0.0537 | **0.3479** | 0.2105 | 0.5748 |
| A64 | A64. | Unspecified sexually transmitted disease | 6612 | 0.0180 | 0.0540 | **0.3331** | 0.2271 | 0.4886 |
| N85 | N85.8 | Other specified noninflammatory disorders of uterus | 3702 | 0.0170 | 0.0537 | **0.3167** | 0.1872 | 0.5360 |
| N46 | N46.9 | Male infertility, unspecified | 2364 | 0.0161 | 0.0536 | **0.2999** | 0.1524 | 0.5904 |
| N84 | N84.1 | Polyp of cervix uteri | 1403 | 0.0157 | 0.0535 | **0.2932** | 0.1204 | 0.7138 |
| N73 | N73.9 | Female pelvic inflammatory disease, unspecified | 1373 | 0.0138 | 0.0535 | **0.2587** | 0.0992 | 0.6745 |
| L68 | L68.0 | Hirsutism | 1024 | 0.0127 | 0.0535 | **0.2375** | 0.0745 | 0.7569 |
| N83 | N83.20 | Unspecified ovarian cysts | 5025 | 0.0127 | 0.0539 | **0.2361** | 0.1400 | 0.3984 |
| N89 | N89.8 | Other specified noninflammatory disorders of vagina | 4793 | 0.0123 | 0.0539 | **0.2283** | 0.1324 | 0.3937 |
| D25 | D25.9 | Leiomyoma of uterus, unspecified | 4634 | 0.0123 | 0.0539 | **0.2282** | 0.1311 | 0.3972 |
| N90 | N90.7 | Vulvar cyst | 988 | 0.0121 | 0.0534 | **0.2272** | 0.0680 | 0.7594 |
| F90 | F90.9 | Attention-deficit hyperactivity disorder, unspecified type | 11738 | 0.0121 | 0.0548 | **0.2207** | 0.1552 | 0.3139 |
| A74 | A74.9 | Chlamydial infection, unspecified | 1082 | 0.0102 | 0.0535 | **0.1901** | 0.0539 | 0.6713 |
| N76 | N76.0 | Acute vaginitis | 10023 | 0.0098 | 0.0547 | **0.1789** | 0.1171 | 0.2733 |
| E28 | E28.2 | Polycystic ovarian syndrome | 2511 | 0.0088 | 0.0537 | **0.1633** | 0.0669 | 0.3987 |
| N94 | N94.6 | Dysmenorrhea, unspecified | 9013 | 0.0081 | 0.0546 | **0.1484** | 0.0908 | 0.2425 |
| N92 | N92.0 | Excessive and frequent menstruation with regular cycle | 9748 | 0.0081 | 0.0547 | **0.1483** | 0.0925 | 0.2377 |
| N77 | N77.1 | Vaginitis, vulvitis and vulvovaginitis in dis classd elswhr | 1805 | 0.0078 | 0.0536 | **0.1448** | 0.0473 | 0.4436 |
| N72 | N72. | Inflammatory disease of cervix uteri | 1687 | 0.0071 | 0.0536 | **0.1328** | 0.0396 | 0.4452 |
| R87 | R87.619 | Unsp abnormal cytolog findings in specmn from cervix uteri | 6501 | 0.0071 | 0.0542 | **0.1305** | 0.0703 | 0.2422 |
| N80 | N80.9 | Endometriosis, unspecified | 2081 | 0.0067 | 0.0536 | **0.1255** | 0.0409 | 0.3846 |
| N87 | N87.9 | Dysplasia of cervix uteri, unspecified | 2020 | 0.0064 | 0.0536 | **0.1200** | 0.0375 | 0.3839 |
| A56 | A56.00 | Chlamydial infection of lower genitourinary tract, unsp | 1100 | 0.0064 | 0.0535 | **0.1190** | 0.0244 | 0.5801 |
| N97 | N97.9 | Female infertility, unspecified | 1964 | 0.0051 | 0.0536 | **0.0950** | 0.0252 | 0.3578 |
| Z30 | Z30.09 | Encounter for oth general cnsl and advice on contraception | 15303 | 0.0050 | 0.0556 | **0.0893** | 0.0551 | 0.1446 |
| N91 | N91.2 | Amenorrhea, unspecified | 2649 | 0.0042 | 0.0537 | **0.0773** | 0.0218 | 0.2740 |
| N93 | N93.9 | Abnormal uterine and vaginal bleeding, unspecified | 5282 | 0.0042 | 0.0541 | **0.0770** | 0.0315 | 0.1884 |
| Z32 | Z32.02 | Encounter for pregnancy test, result negative | 6339 | 0.0024 | 0.0543 | **0.0436** | 0.0147 | 0.1289 |
| Z33 | Z33.1 | Pregnant state, incidental | 3827 | 0.0010 | 0.0539 | **0.0194** | 0.0024 | 0.1585 |

**TABLE 2:** MULTIVARIATE MODEL FOR COVID-19 DEATH BASED UPON PRE-EXISTING CONDITIONS. *Most Common ICD-10 Code and Description for Each Category Diagnosis **Ranked in Descending Order by Contribution to the Logit Function

| RootICD-10 | ICD-10 Code* | ICD-10 Description* | Cases | Coefficient | Std Error | Z_Score | P_Value | Lower CI | Higher CI | Coeff_RR** |
| --- | --- | --- | --- | --- | --- | --- | --- | --- | --- | --- |
| I10 | I10. | Essential (primary) hypertension | 208817 | 0.1934 | 0.0075 | 25.8300 | 0.0000 | 0.1787 | 0.2081 | 0.7424 |
| J64 | J64. | Unspecified pneumoconiosis | 107 | 0.2495 | 0.1007 | 2.4800 | 0.0130 | 0.0521 | 0.4468 | 0.6695 |
| F03 | F03.90 | Unspecified dementia without behavioral disturbance | 13895 | 0.1619 | 0.0076 | 21.3500 | 0.0000 | 0.1471 | 0.1768 | 0.6683 |
| C93 | C93.10 | Chronic myelomonocytic leukemia not achieve remission | 122 | 0.2194 | 0.0815 | 2.6900 | 0.0070 | 0.0597 | 0.3791 | 0.6583 |
| C24 | C24.1 | Malignant neoplasm of ampulla of Vater | 122 | 0.2489 | 0.1006 | 2.4700 | 0.0130 | 0.0517 | 0.4461 | 0.6318 |
| C71 | C71.9 | Malignant neoplasm of brain, unspecified | 557 | 0.3400 | 0.0786 | 4.3300 | 0.0000 | 0.1860 | 0.4941 | 0.5782 |
| C91 | C91.10 | Chronic lymphocytic leuk of B-cell type not achieve remis | 1854 | 0.2492 | 0.0356 | 7.0000 | 0.0000 | 0.1794 | 0.3189 | 0.5362 |
| G70 | G70.00 | Myasthenia gravis without (acute) exacerbation | 745 | 0.2897 | 0.0625 | 4.6300 | 0.0000 | 0.1671 | 0.4122 | 0.5229 |
| D46 | D46.9 | Myelodysplastic syndrome, unspecified | 755 | 0.1633 | 0.0333 | 4.9000 | 0.0000 | 0.0980 | 0.2286 | 0.4858 |
| L12 | L12.0 | Bullous pemphigoid | 334 | 0.1955 | 0.0703 | 2.7800 | 0.0050 | 0.0577 | 0.3334 | 0.4425 |
| C95 | C95.90 | Leukemia, unspecified not having achieved remission | 609 | 0.1530 | 0.0476 | 3.2200 | 0.0010 | 0.0598 | 0.2463 | 0.4055 |
| N18 | N18.9 | Chronic kidney disease, unspecified | 48428 | 0.1663 | 0.0105 | 15.8500 | 0.0000 | 0.1457 | 0.1868 | 0.3998 |
| G20 | G20. | Parkinson's disease | 5105 | 0.1656 | 0.0196 | 8.4500 | 0.0000 | 0.1272 | 0.2040 | 0.3912 |
| C78 | C78.7 | Secondary malig neoplasm of liver and intrahepatic bile duct | 1305 | 0.1545 | 0.0345 | 4.4700 | 0.0000 | 0.0868 | 0.2222 | 0.3834 |
| G30 | G30.9 | Alzheimer's disease, unspecified | 4348 | 0.0866 | 0.0107 | 8.0700 | 0.0000 | 0.0655 | 0.1076 | 0.3712 |
| J84 | J84.10 | Pulmonary fibrosis, unspecified | 7398 | 0.2074 | 0.0225 | 9.2100 | 0.0000 | 0.1633 | 0.2516 | 0.3634 |
| C85 | C85.80 | Oth types of non-Hodgkin lymphoma, unspecified site | 2228 | 0.2301 | 0.0451 | 5.1000 | 0.0000 | 0.1417 | 0.3185 | 0.3588 |
| R18 | R18.8 | Other ascites | 2408 | 0.1669 | 0.0335 | 4.9800 | 0.0000 | 0.1013 | 0.2325 | 0.3367 |
| G11 | G11.1 | Early-onset cerebellar ataxia | 586 | 0.1685 | 0.0640 | 2.6300 | 0.0080 | 0.0431 | 0.2939 | 0.3292 |
| C34 | C34.90 | Malignant neoplasm of unsp part of unsp bronchus or lung | 3805 | 0.1404 | 0.0272 | 5.1500 | 0.0000 | 0.0870 | 0.1938 | 0.3083 |
| Z66 | Z66. | Do not resuscitate | 6448 | 0.0869 | 0.0105 | 8.2600 | 0.0000 | 0.0663 | 0.1075 | 0.3030 |
| F02 | F02.80 | Dementia in oth diseases classd elswhr w/o behavrl disturb | 8684 | 0.0775 | 0.0101 | 7.7100 | 0.0000 | 0.0578 | 0.0972 | 0.3028 |
| R26 | R26.9 | Unspecified abnormalities of gait and mobility | 67067 | 0.1327 | 0.0091 | 14.5100 | 0.0000 | 0.1148 | 0.1506 | 0.2996 |
| C79 | C79.51 | Secondary malignant neoplasm of bone | 2724 | 0.1445 | 0.0299 | 4.8300 | 0.0000 | 0.0859 | 0.2031 | 0.2994 |
| G12 | G12.21 | Amyotrophic lateral sclerosis | 685 | 0.2008 | 0.0842 | 2.3900 | 0.0170 | 0.0359 | 0.3658 | 0.2954 |
| C22 | C22.0 | Liver cell carcinoma | 1150 | 0.1762 | 0.0596 | 2.9600 | 0.0030 | 0.0594 | 0.2929 | 0.2889 |
| D02 | D02.20 | Carcinoma in situ of unspecified bronchus and lung | 2023 | 0.1246 | 0.0342 | 3.6500 | 0.0000 | 0.0577 | 0.1915 | 0.2875 |
| C18 | C18.9 | Malignant neoplasm of colon, unspecified | 4296 | 0.1874 | 0.0333 | 5.6300 | 0.0000 | 0.1221 | 0.2526 | 0.2845 |
| R64 | R64. | Cachexia | 797 | 0.1066 | 0.0384 | 2.7800 | 0.0060 | 0.0313 | 0.1818 | 0.2820 |
| N40 | N40.0 | Benign prostatic hyperplasia without lower urinary tract symp | 85767 | 0.1412 | 0.0103 | 13.6600 | 0.0000 | 0.1209 | 0.1615 | 0.2615 |
| C90 | C90.00 | Multiple myeloma not having achieved remission | 1041 | 0.1577 | 0.0595 | 2.6500 | 0.0080 | 0.0411 | 0.2743 | 0.2591 |
| I25 | I25.10 | Atherosclerotic heart disease of native coronary artery w/o ang pctrs | 77259 | 0.1158 | 0.0095 | 12.1900 | 0.0000 | 0.0972 | 0.1344 | 0.2557 |
| Z96 | Z96.1 | Presence of intraocular lens | 58557 | 0.1228 | 0.0101 | 12.2200 | 0.0000 | 0.1031 | 0.1425 | 0.2537 |
| E11 | E11.9 | Type 2 diabetes mellitus without complications | 114394 | 0.1488 | 0.0118 | 12.5700 | 0.0000 | 0.1256 | 0.1720 | 0.2393 |
| Z74 | Z74.09 | Other reduced mobility | 17940 | 0.0917 | 0.0102 | 8.9500 | 0.0000 | 0.0716 | 0.1117 | 0.2368 |
| L89 | L89.90 | Pressure ulcer of unspecified site, unspecified stage | 8092 | 0.0834 | 0.0130 | 6.4000 | 0.0000 | 0.0579 | 0.1090 | 0.2350 |
| C32 | C32.9 | Malignant neoplasm of larynx, unspecified | 930 | 0.1421 | 0.0653 | 2.1700 | 0.0300 | 0.0140 | 0.2701 | 0.2291 |
| J61 | J61. | Pneumoconiosis due to asbestos and other mineral fibers | 1189 | 0.1202 | 0.0467 | 2.5700 | 0.0100 | 0.0287 | 0.2116 | 0.2271 |
| R54 | R54. | Age-related physical debility | 11357 | 0.0696 | 0.0096 | 7.2600 | 0.0000 | 0.0508 | 0.0884 | 0.2160 |
| J44 | J44.9 | Chronic obstructive pulmonary disease, unspecified | 63653 | 0.1281 | 0.0121 | 10.5500 | 0.0000 | 0.1043 | 0.1519 | 0.2160 |
| I48 | I48.91 | Unspecified atrial fibrillation | 35951 | 0.1048 | 0.0114 | 9.1600 | 0.0000 | 0.0824 | 0.1272 | 0.2035 |
| I96 | I96. | Gangrene, not elsewhere classified | 2079 | 0.0857 | 0.0322 | 2.6600 | 0.0080 | 0.0225 | 0.1488 | 0.1925 |
| I37 | I37.0 | Nonrheumatic pulmonary valve stenosis | 1552 | 0.1488 | 0.0670 | 2.2200 | 0.0260 | 0.0175 | 0.2800 | 0.1912 |
| E78 | E78.5 | Hyperlipidemia, unspecified | 223873 | 0.0921 | 0.0127 | 7.2500 | 0.0000 | 0.0672 | 0.1170 | 0.1897 |
| I71 | I71.4 | Abdominal aortic aneurysm, without rupture | 12513 | 0.1183 | 0.0192 | 6.1800 | 0.0000 | 0.0808 | 0.1559 | 0.1896 |
| I50 | I50.9 | Heart failure, unspecified | 36427 | 0.0771 | 0.0102 | 7.5800 | 0.0000 | 0.0572 | 0.0971 | 0.1863 |
| H25 | H25.13 | Age-related nuclear cataract, bilateral | 127537 | 0.1014 | 0.0129 | 7.8700 | 0.0000 | 0.0762 | 0.1267 | 0.1799 |
| I85 | I85.00 | Esophageal varices without bleeding | 2480 | 0.1587 | 0.0712 | 2.2300 | 0.0260 | 0.0192 | 0.2983 | 0.1764 |
| J96 | J96.01 | Acute respiratory failure with hypoxia | 13350 | 0.0753 | 0.0135 | 5.5600 | 0.0000 | 0.0488 | 0.1019 | 0.1749 |
| N19 | N19. | Unspecified kidney failure | 7383 | 0.0749 | 0.0170 | 4.4200 | 0.0000 | 0.0417 | 0.1082 | 0.1702 |
| J47 | J47.9 | Bronchiectasis, uncomplicated | 1749 | 0.0978 | 0.0454 | 2.1600 | 0.0310 | 0.0089 | 0.1867 | 0.1656 |
| M81 | M81.0 | Age-related osteoporosis w/o current pathological fracture | 10016 | 0.1163 | 0.0249 | 4.6700 | 0.0000 | 0.0675 | 0.1652 | 0.1645 |
| D04 | D04.9 | Carcinoma in situ of skin, unspecified | 3845 | 0.1033 | 0.0345 | 3.0000 | 0.0030 | 0.0357 | 0.1708 | 0.1639 |
| H27 | H27.8 | Other specified disorders of lens | 6825 | 0.0739 | 0.0173 | 4.2800 | 0.0000 | 0.0401 | 0.1078 | 0.1638 |
| H90 | H90.3 | Sensorineural hearing loss, bilateral | 118138 | 0.1200 | 0.0145 | 8.2600 | 0.0000 | 0.0915 | 0.1485 | 0.1634 |
| E03 | E03.9 | Hypothyroidism, unspecified | 35891 | 0.1812 | 0.0244 | 7.4300 | 0.0000 | 0.1334 | 0.2290 | 0.1631 |
| D64 | D64.9 | Anemia, unspecified | 62258 | 0.0916 | 0.0122 | 7.5300 | 0.0000 | 0.0677 | 0.1154 | 0.1576 |
| B02 | B02.9 | Zoster without complications | 16479 | 0.1667 | 0.0329 | 5.0700 | 0.0000 | 0.1022 | 0.2312 | 0.1520 |
| N25 | N25.81 | Secondary hyperparathyroidism of renal origin | 6475 | 0.0635 | 0.0191 | 3.3300 | 0.0010 | 0.0261 | 0.1008 | 0.1478 |
| I73 | I73.9 | Peripheral vascular disease, unspecified | 31350 | 0.0715 | 0.0114 | 6.2800 | 0.0000 | 0.0492 | 0.0938 | 0.1469 |
| H61 | H61.23 | Impacted cerumen, bilateral | 58221 | 0.1234 | 0.0162 | 7.6000 | 0.0000 | 0.0916 | 0.1552 | 0.1453 |
| Z43 | Z43.3 | Encounter for attention to colostomy | 2992 | 0.0951 | 0.0412 | 2.3100 | 0.0210 | 0.0144 | 0.1758 | 0.1428 |
| H91 | H91.90 | Unspecified hearing loss, unspecified ear | 71156 | 0.1438 | 0.0204 | 7.0600 | 0.0000 | 0.1039 | 0.1838 | 0.1408 |
| H26 | H26.9 | Unspecified cataract | 60786 | 0.0719 | 0.0110 | 6.5600 | 0.0000 | 0.0504 | 0.0933 | 0.1370 |
| H35 | H35.31 | Nonexudative age-related macular degeneration | 60393 | 0.0900 | 0.0132 | 6.8200 | 0.0000 | 0.0641 | 0.1158 | 0.1344 |
| C67 | C67.9 | Malignant neoplasm of bladder, unspecified | 4135 | 0.0867 | 0.0338 | 2.5600 | 0.0100 | 0.0203 | 0.1530 | 0.1324 |
| Z95 | Z95.1 | Presence of aortocoronary bypass graft | 34407 | 0.0659 | 0.0121 | 5.4600 | 0.0000 | 0.0422 | 0.0896 | 0.1317 |
| R62 | R62.7 | Adult failure to thrive | 3414 | 0.0395 | 0.0150 | 2.6400 | 0.0080 | 0.0102 | 0.0689 | 0.1295 |
| J90 | J90. | Pleural effusion, not elsewhere classified | 5673 | 0.0519 | 0.0175 | 2.9600 | 0.0030 | 0.0176 | 0.0862 | 0.1257 |
| R63 | R63.4 | Abnormal weight loss | 34666 | 0.1042 | 0.0190 | 5.4700 | 0.0000 | 0.0669 | 0.1415 | 0.1253 |
| J43 | J43.9 | Emphysema, unspecified | 9950 | 0.0875 | 0.0256 | 3.4100 | 0.0010 | 0.0372 | 0.1377 | 0.1241 |
| L97 | L97.509 | Non-pressure chronic ulcer oth prt unsp foot w unsp severity | 13950 | 0.0648 | 0.0189 | 3.4300 | 0.0010 | 0.0278 | 0.1019 | 0.1181 |
| R60 | R60.0 | Localized edema | 61977 | 0.0741 | 0.0128 | 5.7900 | 0.0000 | 0.0490 | 0.0992 | 0.1161 |
| C61 | C61. | Malignant neoplasm of prostate | 19707 | 0.0974 | 0.0262 | 3.7100 | 0.0000 | 0.0460 | 0.1489 | 0.1106 |
| H54 | H54.7 | Unspecified visual loss | 25803 | 0.0907 | 0.0203 | 4.4800 | 0.0000 | 0.0510 | 0.1303 | 0.1098 |
| K74 | K74.60 | Unspecified cirrhosis of liver | 8439 | 0.1148 | 0.0497 | 2.3100 | 0.0210 | 0.0175 | 0.2122 | 0.1076 |
| D63 | D63.1 | Anemia in chronic kidney disease | 14823 | 0.0421 | 0.0124 | 3.3900 | 0.0010 | 0.0177 | 0.0665 | 0.1073 |
| N47 | N47.6 | Balanoposthitis | 6273 | 0.0979 | 0.0421 | 2.3300 | 0.0200 | 0.0155 | 0.1804 | 0.1054 |
| M10 | M10.9 | Gout, unspecified | 32778 | 0.1014 | 0.0223 | 4.5400 | 0.0000 | 0.0576 | 0.1452 | 0.1035 |
| I67 | I67.89 | Other cerebrovascular disease | 16692 | 0.0534 | 0.0157 | 3.4000 | 0.0010 | 0.0226 | 0.0842 | 0.1021 |
| I70 | I70.0 | Atherosclerosis of aorta | 19489 | 0.0537 | 0.0147 | 3.6400 | 0.0000 | 0.0248 | 0.0825 | 0.1000 |
| C44 | C44.91 | Basal cell carcinoma of skin, unspecified | 21770 | 0.0769 | 0.0250 | 3.0800 | 0.0020 | 0.0280 | 0.1259 | 0.0906 |
| I63 | I63.9 | Cerebral infarction, unspecified | 18075 | 0.0507 | 0.0173 | 2.9400 | 0.0030 | 0.0169 | 0.0845 | 0.0905 |
| T82 | T82.818A | Embolism due to vascular prosth dev/grft, initial encounter | 6593 | 0.0462 | 0.0232 | 1.9900 | 0.0470 | 0.0006 | 0.0917 | 0.0866 |
| M15 | M15.9 | Polyosteoarthritis, unspecified | 70950 | 0.0799 | 0.0191 | 4.1800 | 0.0000 | 0.0424 | 0.1173 | 0.0826 |
| J18 | J18.9 | Pneumonia, unspecified organism | 32422 | 0.0512 | 0.0155 | 3.3000 | 0.0010 | 0.0208 | 0.0817 | 0.0796 |
| Z65 | Z65.9 | Problem related to unspecified psychosocial circumstances | 127694 | 0.0833 | 0.0209 | 3.9800 | 0.0000 | 0.0423 | 0.1243 | 0.0730 |
| I65 | I65.29 | Occlusion and stenosis of unspecified carotid artery | 16848 | 0.0413 | 0.0162 | 2.5500 | 0.0110 | 0.0096 | 0.0729 | 0.0717 |
| R01 | R01.1 | Cardiac murmur, unspecified | 14466 | 0.0708 | 0.0309 | 2.2900 | 0.0220 | 0.0102 | 0.1313 | 0.0715 |
| R33 | R33.9 | Retention of urine, unspecified | 20323 | 0.0408 | 0.0164 | 2.4900 | 0.0130 | 0.0086 | 0.0729 | 0.0694 |
| R32 | R32. | Unspecified urinary incontinence | 18819 | 0.0441 | 0.0192 | 2.2900 | 0.0220 | 0.0064 | 0.0817 | 0.0650 |
| R80 | R80.3 | Bence Jones proteinuria | 13651 | 0.0517 | 0.0262 | 1.9700 | 0.0490 | 0.0003 | 0.1030 | 0.0646 |
| N17 | N17.9 | Acute kidney failure, unspecified | 31785 | 0.0282 | 0.0123 | 2.2900 | 0.0220 | 0.0041 | 0.0523 | 0.0604 |
| R97 | R97.2 | Elevated prostate specific antigen [PSA] | 36803 | 0.0617 | 0.0248 | 2.4900 | 0.0130 | 0.0131 | 0.1103 | 0.0598 |
| H02 | H02.839 | Dermatochalasis of unspecified eye, unspecified eyelid | 29211 | 0.0526 | 0.0216 | 2.4300 | 0.0150 | 0.0102 | 0.0950 | 0.0580 |
| L57 | L57.0 | Actinic keratosis | 46813 | 0.0494 | 0.0226 | 2.1900 | 0.0290 | 0.0051 | 0.0937 | 0.0505 |
| E87 | E87.6 | Hypokalemia | 54353 | 0.0312 | 0.0138 | 2.2600 | 0.0240 | 0.0041 | 0.0583 | 0.0492 |
| R91 | R91.8 | Other nonspecific abnormal finding of lung field | 37585 | -0.0542 | 0.0251 | -2.1600 | 0.0310 | -0.1035 | -0.0049 | -0.0523 |
| I51 | I51.7 | Cardiomegaly | 24480 | -0.0507 | 0.0225 | -2.2500 | 0.0240 | -0.0948 | -0.0066 | -0.0592 |
| R31 | R31.9 | Hematuria, unspecified | 42562 | -0.0824 | 0.0309 | -2.6700 | 0.0080 | -0.1430 | -0.0219 | -0.0600 |
| R78 | R78.89 | Finding of oth substances, not normally found in blood | 32503 | -0.0692 | 0.0268 | -2.5800 | 0.0100 | -0.1217 | -0.0166 | -0.0624 |
| Z09 | Z09. | Encntr for f/u exam aft trtmt for cond oth than malig neoplms | 55214 | -0.0987 | 0.0319 | -3.0900 | 0.0020 | -0.1612 | -0.0361 | -0.0660 |
| R42 | R42. | Dizziness and giddiness | 67634 | -0.0950 | 0.0282 | -3.3700 | 0.0010 | -0.1503 | -0.0398 | -0.0677 |
| R55 | R55. | Syncope and collapse | 30812 | -0.0755 | 0.0277 | -2.7200 | 0.0070 | -0.1298 | -0.0211 | -0.0680 |
| M62 | M62.81 | Muscle weakness (generalized) | 71400 | -0.0695 | 0.0206 | -3.3700 | 0.0010 | -0.1099 | -0.0291 | -0.0696 |
| Z86 | Z86.010 | Personal history of colonic polyps | 103774 | -0.0745 | 0.0197 | -3.7700 | 0.0000 | -0.1132 | -0.0358 | -0.0697 |
| N13 | N13.30 | Unspecified hydronephrosis | 13082 | -0.0708 | 0.0338 | -2.1000 | 0.0360 | -0.1369 | -0.0046 | -0.0727 |
| I12 | I12.9 | Hypertensive chronic kidney disease w stg 1-4/unsp chr kdny | 26422 | -0.0329 | 0.0131 | -2.5200 | 0.0120 | -0.0585 | -0.0073 | -0.0762 |
| E83 | E83.42 | Hypomagnesemia | 22641 | -0.0670 | 0.0234 | -2.8700 | 0.0040 | -0.1128 | -0.0212 | -0.0792 |
| L03 | L03.90 | Cellulitis, unspecified | 70833 | -0.1097 | 0.0282 | -3.8900 | 0.0000 | -0.1651 | -0.0544 | -0.0797 |
| L98 | L98.9 | Disorder of the skin and subcutaneous tissue, unspecified | 55257 | -0.1296 | 0.0345 | -3.7600 | 0.0000 | -0.1971 | -0.0621 | -0.0801 |
| F06 | F06.30 | Mood disorder due to known physiological condition, unsp | 31408 | -0.0931 | 0.0291 | -3.2000 | 0.0010 | -0.1502 | -0.0360 | -0.0816 |
| G93 | G93.3 | Postviral fatigue syndrome | 39656 | -0.0957 | 0.0253 | -3.7800 | 0.0000 | -0.1453 | -0.0461 | -0.0881 |
| E86 | E86.0 | Dehydration | 28085 | -0.0714 | 0.0210 | -3.4000 | 0.0010 | -0.1126 | -0.0302 | -0.0899 |
| Z79 | Z79.899 | Other long term (current) drug therapy | 147374 | -0.0736 | 0.0162 | -4.5500 | 0.0000 | -0.1054 | -0.0419 | -0.0915 |
| D50 | D50.9 | Iron deficiency anemia, unspecified | 31287 | -0.0753 | 0.0208 | -3.6200 | 0.0000 | -0.1160 | -0.0345 | -0.0919 |
| Z91 | Z91.19 | Patient's noncompliance w oth medical treatment and regimen | 64332 | -0.1130 | 0.0250 | -4.5200 | 0.0000 | -0.1620 | -0.0640 | -0.0932 |
| K56 | K56.60 | Unspecified intestinal obstruction | 8535 | -0.0953 | 0.0406 | -2.3500 | 0.0190 | -0.1748 | -0.0159 | -0.0966 |
| M20 | M20.40 | Other hammer toe(s) (acquired), unspecified foot | 43692 | -0.1552 | 0.0345 | -4.5000 | 0.0000 | -0.2228 | -0.0875 | -0.1006 |
| I47 | I47.1 | Supraventricular tachycardia | 12549 | -0.1055 | 0.0346 | -3.0500 | 0.0020 | -0.1732 | -0.0377 | -0.1061 |
| M19 | M19.90 | Unspecified osteoarthritis, unspecified site | 89625 | -0.1818 | 0.0320 | -5.6800 | 0.0000 | -0.2445 | -0.1190 | -0.1095 |
| B95 | B95.61 | Methicillin suscep staph infct causing dis classd elswhr | 13437 | -0.1003 | 0.0321 | -3.1300 | 0.0020 | -0.1631 | -0.0374 | -0.1130 |
| R25 | R25.2 | Cramp and spasm | 28206 | -0.1680 | 0.0389 | -4.3200 | 0.0000 | -0.2443 | -0.0918 | -0.1133 |
| F05 | F05. | Delirium due to known physiological condition | 3612 | -0.0458 | 0.0203 | -2.2600 | 0.0240 | -0.0855 | -0.0061 | -0.1148 |
| J98 | J98.4 | Other disorders of lung | 35483 | -0.1159 | 0.0247 | -4.6800 | 0.0000 | -0.1644 | -0.0674 | -0.1154 |
| S22 | S22.39XA | Fracture of one rib, unsp side, init for clos fx | 10436 | -0.1438 | 0.0473 | -3.0400 | 0.0020 | -0.2366 | -0.0511 | -0.1165 |
| K31 | K31.89 | Other diseases of stomach and duodenum | 15241 | -0.1706 | 0.0496 | -3.4400 | 0.0010 | -0.2679 | -0.0734 | -0.1169 |
| G46 | G46.4 | Cerebellar stroke syndrome | 4838 | -0.0887 | 0.0379 | -2.3400 | 0.0190 | -0.1631 | -0.0144 | -0.1205 |
| I11 | I11.9 | Hypertensive heart disease without heart failure | 28332 | -0.0920 | 0.0190 | -4.8500 | 0.0000 | -0.1291 | -0.0548 | -0.1219 |
| I13 | I13.0 | Hyp hrt & chr kdny dis w hrt fail and stg 1-4/unsp chr kdny | 9823 | -0.0454 | 0.0133 | -3.4100 | 0.0010 | -0.0716 | -0.0193 | -0.1239 |
| Z92 | Z92.3 | Personal history of irradiation | 8961 | -0.1416 | 0.0468 | -3.0200 | 0.0030 | -0.2334 | -0.0498 | -0.1287 |
| H43 | H43.819 | Vitreous degeneration, unspecified eye | 38102 | -0.1599 | 0.0280 | -5.7000 | 0.0000 | -0.2148 | -0.1049 | -0.1304 |
| Z48 | Z48.02 | Encounter for removal of sutures | 78659 | -0.1961 | 0.0303 | -6.4800 | 0.0000 | -0.2554 | -0.1368 | -0.1306 |
| G81 | G81.90 | Hemiplegia, unspecified affecting unspecified side | 3445 | -0.0999 | 0.0456 | -2.1900 | 0.0280 | -0.1893 | -0.0105 | -0.1327 |
| Y83 | Y83.1 | Implnt of artif int dev cause abn react/compl, w/o misadvnt | 11053 | -0.1500 | 0.0426 | -3.5200 | 0.0000 | -0.2335 | -0.0666 | -0.1394 |
| D62 | D62. | Acute posthemorrhagic anemia | 6406 | -0.1002 | 0.0306 | -3.2800 | 0.0010 | -0.1601 | -0.0403 | -0.1441 |
| I20 | I20.8 | Other forms of angina pectoris | 31188 | -0.1400 | 0.0241 | -5.8100 | 0.0000 | -0.1872 | -0.0928 | -0.1445 |
| R06 | R06.02 | Shortness of breath | 118999 | -0.1827 | 0.0247 | -7.3900 | 0.0000 | -0.2311 | -0.1342 | -0.1459 |
| G62 | G62.9 | Polyneuropathy, unspecified | 14108 | -0.1832 | 0.0409 | -4.4800 | 0.0000 | -0.2635 | -0.1030 | -0.1496 |
| Z13 | Z13.6 | Encounter for screening for cardiovascular disorders | 237211 | -0.1886 | 0.0286 | -6.5900 | 0.0000 | -0.2447 | -0.1325 | -0.1519 |
| S91 | S91.309A | Unspecified open wound, unspecified foot, initial encounter | 11184 | -0.1519 | 0.0374 | -4.0600 | 0.0000 | -0.2253 | -0.0786 | -0.1587 |
| R39 | R39.15 | Urgency of urination | 29413 | -0.2332 | 0.0372 | -6.2700 | 0.0000 | -0.3061 | -0.1604 | -0.1648 |
| Z98 | Z98.890 | Other specified postprocedural states | 92911 | -0.2348 | 0.0272 | -8.6400 | 0.0000 | -0.2881 | -0.1815 | -0.1663 |
| T83 | T83.098A | Mech compl of other urinary catheter, initial encounter | 3957 | -0.1205 | 0.0408 | -2.9500 | 0.0030 | -0.2004 | -0.0406 | -0.1685 |
| H52 | H52.4 | Presbyopia | 192666 | -0.1513 | 0.0212 | -7.1400 | 0.0000 | -0.1928 | -0.1097 | -0.1731 |
| I16 | I16.0 | Hypertensive urgency | 5096 | -0.2349 | 0.0548 | -4.2900 | 0.0000 | -0.3423 | -0.1275 | -0.2227 |
| J16 | J16.8 | Pneumonia due to other specified infectious organisms | 1240 | -0.1815 | 0.0750 | -2.4200 | 0.0160 | -0.3285 | -0.0345 | -0.2321 |
| Z23 | Z23. | Encounter for immunization | 260232 | -0.3022 | 0.0320 | -9.4500 | 0.0000 | -0.3649 | -0.2395 | -0.2325 |
| G21 | G21.11 | Neuroleptic induced parkinsonism | 2043 | -0.1384 | 0.0412 | -3.3600 | 0.0010 | -0.2193 | -0.0576 | -0.2494 |
| I97 | I97.710 | Intraoperative cardiac arrest during cardiac surgery | 2682 | -0.2555 | 0.0660 | -3.8700 | 0.0000 | -0.3849 | -0.1260 | -0.2650 |
| Z16 | Z16.11 | Resistance to penicillins | 2546 | -0.2529 | 0.0682 | -3.7100 | 0.0000 | -0.3866 | -0.1192 | -0.2690 |
| T87 | T87.89 | Other complications of amputation stump | 1707 | -0.2549 | 0.0842 | -3.0300 | 0.0020 | -0.4199 | -0.0899 | -0.2772 |
| W05 | W05.0XXA | Fall from non-moving wheelchair, initial encounter | 660 | -0.1834 | 0.0706 | -2.6000 | 0.0090 | -0.3219 | -0.0450 | -0.3132 |

Tables 1 and 2 show that this approach provides much more clinical information than the indices. Table 1 ranks the category diagnoses by their RR on univariate analysis. Degenerative neurologic disease and severe functional disability are prominently represented within the top 20 most influential conditions. This observation is significant because they are not given much weight in comorbidity indices. On the other hand, only one solid tumor (carcinoma of the ampulla of Vater) appears within the top 20. On the other end of the scale, several functional diagnoses, gynecologic disorders, and sexually transmitted diseases reduced the risk of death. Table 2 shows the ICD-10 codes that were independently predictive of death rank ordered by their contribution to the logit when present. Many high-risk conditions became protective when adjusted for the effects of others. Hypertension was the most important independent risk factor for death and represented a greater threat than coronary artery disease. Degenerative neurologic diseases were prominently represented at the top of the list, while malignancies comprised the bulk of high-risk conditions.

## DISCUSSION

We propose a novel method for handling pre-existing conditions in COVID prediction models that has several theoretical advantages over conventional comorbidity indices. Table 3 compares the attributes of the standard comorbidity indices and PDeathDx. Unlike a standard index, our approach creates a different model for every disease. The outcome can be changed by re-defining a target group. Univariate statistical analysis is used to assign condition weights. The method can be applied to any type of categorical predictor including medications and procedures. It handles rare as well as protective conditions. In addition to identifying high-risk codes, the output includes a probability for each pattern of conditions. It can therefore detect circumstances where disease combinations are problematic. Finally, the model can be used to return actional information to providers. Instead of total or condition scores, this information includes the predicted probability of the outcome, the specific diagnoses contributing to the prediction, and their rank order of importance. Such information may prompt the provider to explore a mechanism of injury or an intervention to mitigate the risk.

**TABLE 3:** COMPARISON OF METHODS FOR HANDLING COMORBIDITIES.

|  | **Charleson** | **Elixhauser** | **P(Death)Dx** |
| --- | --- | --- | --- |
| Diagnosis-specific | No | No | Yes |
| Includes all relevant diagnosis | No | No | Yes |
| Number of comorbidities | 19 | 30 | — |
| Weighting | Arbitrary | None | Actual risk |
| Includes protective factors | No | Yes | Yes |
| Handles rare conditions | No | No | Yes |
| Effect of each comorbidity adjusted for others | No | No | Yes |
| Identifies all comorbid patterns and associated risks | No | No | Yes |
| Setting of extracted diagnostic codes | Inpatient only | Inpatient only | Inpatient/outpatient |

However, PDeathDx requires programming at a high level, takes more time, and uses more computing resources. The first hurdle is the systematic search for candidate diagnoses among millions of entries in the electronic record and performing screening statistical tests on thousands of root diagnoses. The second is creating a diagnostic array in which each patient is given a score for every significant diagnosis experienced by the cohort. The table must then be transferred to a statistical program capable of handling hundreds of predictors and large number of rows.

PDeathDx provided powerful discrimination between COVID-19 patients who died and survived, out-performed the other comorbidity indices, had an ROC area equivalent to that generated by age, and remains the second most powerful predictor of all those that we have examined thus far. The most influential category conditions included entries not usually included in comorbidity indices. This observation suggests that it is risky to pick comorbidities for analysis without a systematic review of all those experienced by the cohort. Moreover, poor prognoses were assigned to those with dementia, degenerative neurological diseases, and severe disabilities – conditions not usually associated with cardiorespiratory injury or impaired immunity. These disorders are often associated with a poor quality of life that dictates a conservative approach to treatment. If this decision is common for COVID-19 patients, then death may be more indicative of the patient’s baseline condition than severity of illness. In that event, this outcome may not be suitable for studies of interventions. Finally, multivariate analysis showed that some high-risk conditions became protective when adjusted for the effect of other components of the model. It would have been a mistake to treat the effect of these conditions as independent and additive.

We did not include age in our model for PDeathDx for 3 reasons. First, it was intended to represent pre-existing conditions in an overarching model containing multiple domains. One domain was comprised of demographic characteristics including age at diagnosis. Next, we did not want age to displace the diagnoses highly correlated with age in the model. To understand the mechanisms that lead to a fatal outcome, explanatory variables should take precedence over disease markers. If age is modelled at all, it should be for a residual effect once causal factors have been included. Finally, age-based models do not support decision-making at the point of care. For example, a physician might feel comfortable withdrawing care if the patient had Alzheimer’s disease – but not simply because the patient was old. Clinicians make recommendations based upon an assessment of the underlying conditions. It is the patient’s prerogative to decide what is appropriate based upon age and quality of life.

We included all diagnoses in the medical record because some conditions have an effect over the patient’s lifetime even if they have not been recently active. Examples include intravenous drug use or sickle cell trait. We also wanted to test our computing resources to see if they were capable of handling problems of this size. Finally, to develop the most efficient model, it is reasonable to start with a comprehensive list of diagnoses and work backwards. That strategy allows one to determine if time-limited sampling compromises the ability of PDeathDx to discriminate between survivors and non-survivors. On the other hand, starting with a fixed time frame for diagnoses is arbitrary and provides no information about the benefits of more remote data. Fortunately, our computing resources were more than sufficient to handle all pre-existing conditions.

The major limitation of our approach is that it handles all pre-existing diagnoses – not just the most recent ones. Thus, a person with chronic renal failure (CRF) who undergoes a transplant and regains normal renal function will still be included in the analysis of CRF. Of course, our conclusions are limited to patients with characteristics like the veteran population. Further studies should be done on other populations and disease states before the method should be widely applied. If validated by others, our method could provide a more robust alternative to comorbidity scores for handling pre-existing conditions in multivariate models.

## Data Availability

All data produced in the present study are available upon reasonable request to the authors.

